# Serological markers and Post COVID-19 Condition (PCC) – A rapid review of the evidence

**DOI:** 10.1101/2023.10.30.23297455

**Authors:** Erin Collins, Elizabeth Philippe, Chris Gravel, Steven Hawken, Marc-André Langlois, Julian Little

**Author notes:** **Correspondence:** Erin Collins, School of Epidemiology and Public Health, Faculty of Medicine, University of Ottawa. Ottawa, Ontario, Canada. 451 Smyth Road, K1H 8M5. Julian Little, School of Epidemiology and Public Health, Faculty of Medicine, University of Ottawa. Ottawa, Ontario, Canada. 451 Smyth Road, K1H 8M5.

## Abstract

**Background:** Post COVID-19 Condition (PCC) is highly heterogeneous, often debilitating, and may last for years after infection. The etiology of PCC remains uncertain. Examination of potential serological markers of PCC, accounting for clinical covariates, may yield emergent pathophysiological insights.

**Methods:** In adherence to PRISMA guidelines, we carried out a rapid review of the literature. We searched Medline and Embase for primary observational studies that compared IgG response in individuals who experienced COVID-19 symptoms persisting ≥12 weeks post-infection with those who did not. We examined relationships between serological markers and PCC status and investigated sources of inter-study variability, such as severity of acute illness, PCC symptoms assessed, and target antigen(s).

**Results:** Of 8,018 unique records, we identified 29 as being eligible for inclusion in synthesis. Definitions of PCC varied. In studies that reported anti-nucleocapsid (N) IgG (n=10 studies; n=989 participants in aggregate), full or partial anti-Spike IgG (i.e., the whole trimer, S1 or S2 subgroups, or receptor binding domain, n=19 studies; n=2606 participants), or neutralizing response (n=7 studies; n=1123 participants), we did not find strong evidence to support any difference in serological markers between groups with and without persisting symptoms. However, most studies did not account for severity or level of care required during acute illness, and other potential confounders.

**Conclusions:** Pooling of studies would enable more robust exploration of clinical and serological predictors among diverse populations. However, substantial inter-study variations hamper comparability. Standardized reporting practices would improve the quality, consistency, and comprehension of study findings.

## Introduction

Post COVID-19 Condition (PCC) broadly refers to the persistence of symptoms occurring three months or longer post-infection [1–3]. PCC is highly heterogeneous and may manifest as different clusters of symptoms of varying severity and duration [3–7]. While the prevalence of PCC has been found to decrease with increasing months post-infection [8,9], the condition may persist over two years [9,10]. PCC can often have debilitating and wide-ranging impacts, such as diminished quality of life, inability to work or attend school, need for healthcare services, reduced work productivity, and reliance on caregiver support [3,4,10–13]. The etiology of PCC remains uncertain, though several underlying pathophysiological mechanisms, such as cellular damage, inflammatory cytokines, and a hypercoagulable state, are thought to contribute to PCC inception and trajectory [7, 14–17].

Given the complexity of the condition, a diverse range of potential predictors warrant consideration. Older age, female sex, pre-existing conditions (e.g., high BMI, asthma, and diabetes), and severity of acute illness, have frequently been proposed as risk factors for post-acute sequelae [3,4,15,18–20]. Additionally, a number of biomarkers have been investigated but currently, there is no consensus as to whether any characterize PCC [14,16,17].

Investigation of potential serological markers of PCC, accounting for clinical covariates, may yield pathophysiological insights. To date, several observational studies have compared humoral response between groups with and without persistent symptoms, albeit with highly mixed findings. Most of the evidence to date is on adult populations. Given the utility of serological testing to identify past infection, these efforts may illuminate potential differences in antibody detection that are associated with the presence of persisting symptoms, or specific PCC phenotypes [5,12,17,21–30]. Some studies have found that people with PCC are more likely to elicit a robust humoral response, as compared to people with past COVID-19 infection and no PCC, which could result from viral antigen persistence or over-activation of the immune system [24–27]. On the other hand, findings that people with PCC are more prone to non-response, weak response, or early waning of antibodies may indicate impaired functional antiviral response [21, 23, 28–30]. However, investigation of associations between PCC and serological markers are complicated by differences in inclusion criteria, study procedures, serological assays, choice of antibody and target antigen, timing of follow-up for PCC assessment and serological sampling, methods of statistical analysis, and completeness of reporting [3,12,31]. COVID-19 variant and vaccination status may also influence findings [32–36].

We performed a systematic search of the literature to collect and collate serological comparisons between adults with and without persistent symptoms following COVID-19 infection. The aims of this review were to 1) assess relationships between post-infection serological response and PCC, and 2) investigate and report on sources of inter-study heterogeneity.

## Methods

We completed a rapid review [37] of the literature to examine serological results compared between groups with and without persistent symptoms post COVID-19 infection. We reported findings in accordance with the Preferred Reporting Items for Systematic Review and Meta-Analysis (PRISMA) Statement [38] (**Supplementary Materials**), and registered our review in PROSPERO (CRD42023402978). A protocol was not prepared.

### Search strategy and study eligibility

We searched Medline and Embase for reports published between January 1, 2020 – October 22, 2022. We imposed no language restrictions on the search. We used a search strategy with key terms relating to 1) Post COVID-19 Condition, and 2) observational studies (**Supplementary Materials**).

We included records which met the following criteria:

- Primary observational study;
- Language: English, French, or Italian;
- ≥50 participants and ≥75% adults (≥16 years of age) assessed for persistent symptoms ≥12 weeks post COVID-19 onset/diagnosis;
- ≥1 post-acute (≥4 weeks post COVID-19) serology result reported for a) individuals with any persistent symptoms or a persistent symptom(s) of interest (e.g., post-acute fatigue), and compared with results from individuals without any persistent symptoms or a persistent symptom(s) of interest; or b) individuals with varying PCC severity.

Preprints were included so long as other eligibility criteria were met.

### Study selection and data extraction

Records identified using the search strategy were entered into Covidence Systematic Review Software. All abstracts and full texts were screened for potential inclusion by one author (EC) using pre-piloted criteria generated by consensus. A second author (EP) verified 10% of records, until a kappa/interrater agreement > 0.8 was achieved. An extraction file was created by consensus and piloted in Excel 2016. Two reviewers (EC and EP) extracted data and 10% of extractions were verified until kappa >0.8. In the event of a disagreement that could not be resolved by consensus, a third reviewer was available to address (JL).

We extracted study characteristics, PCC description and duration, and serological results. We also extracted variables that may have influenced serological results and/or PCC character and trajectory, such as timing of COVID-19 infection, COVID-19 variants and vaccination status, and potential individual-level confounders we identified a priori (age, sex, level of care (LOC) during acute illness; severity of acute illness; number of acute symptoms; and pre-existing conditions, including diabetes, chronic respiratory illness, cardiac disease, and conditions or medications which may suppress immune function). If COVID-19 variant was not specified, we identified that which prevailed in the host country when participants were infected or recruited [39]. If vaccination status was not recorded, we assumed the study population to be non-vaccinated at time of infection if dates of infection or recruitment preceded mass vaccination efforts in the host country [40]. With relation to LOC, we identified study populations as hospitalized, non-hospitalized, or having “mixed” LOC requirements during acute illness. A population was defined as “mixed” if the proportions of hospitalized and non-hospitalized study participants both exceeded 5%.

### Evaluation of risk of bias

We used the Newcastle-Ottawa quality assessment scale (NOS) for observational studies to evaluate quality and risk of bias, and an adapted scale for cross sectional studies [41]. The NOS scale assigns points based on selection, comparability, and outcome of interest. A maximum of nine points was assigned to cohort and case control studies and cross-sectional studies were scored up to seven points. Two authors (EC and EP) independently assessed risk of bias, and 10% of studies were cross-checked by a second author. In the event of a disagreement that could not be resolved by consensus, a third reviewer was available to resolve any disagreements that could not be resolved by consensus (JL).

### Data synthesis

We compared measures of effect (difference, average, prevalence, or risk) of serological response corresponding to PCC status. Given high inter-study variability, we determined that a meta-analysis of results was not appropriate and instead presented a narrative description of findings. We reported the overall trend in IgG response to SARS-CoV-2 infection, when results among people with persistent symptoms were compared to those without. The trend of each study was classified as a) increase (if ≥ one increase reported), b) decrease (if ≥ one decrease reported), or c) no increase/decrease (if no increase or decrease reported).

Given multiple reports on the same study population, we distinguished between “study population” and “report”, the latter of which refers to each record included in synthesis. We summarized overall associations between serological levels and PCC (as defined by study authors), and sources of inter-study heterogeneity. We presented results stratified by LOC and timing of serological follow-up.

## Results

### Study selection and study population characteristics

After removal of duplicates (n=922), we screened 8,018 abstracts and 2,000 full texts, of which 29 records (23 study populations) met eligibility criteria and were included in synthesis (**Figure 1**). **Table 1** summarizes the characteristics of included studies. Studies were published between March 2021 to September 2022 and sample sizes ranged between 51 and 589. Most study populations were “mixed”, i.e., either hospitalized or non-hospitalized during acute illness (n=14), while five were non-hospitalized and three were hospitalized. One study did not specify level of care (LOC) [42], and was hence excluded from synthesis by LOC status (**Tables S1 and S2**). We captured severity of acute illness in addition to LOC, though the high variety of scales used to assess severity limited inter-study comparability. We also collected any information on number of symptoms during acute illness, given this feature has been found to be predictive of PCC [18], though these data were available for few study populations (n=4).

**Figure 1:**
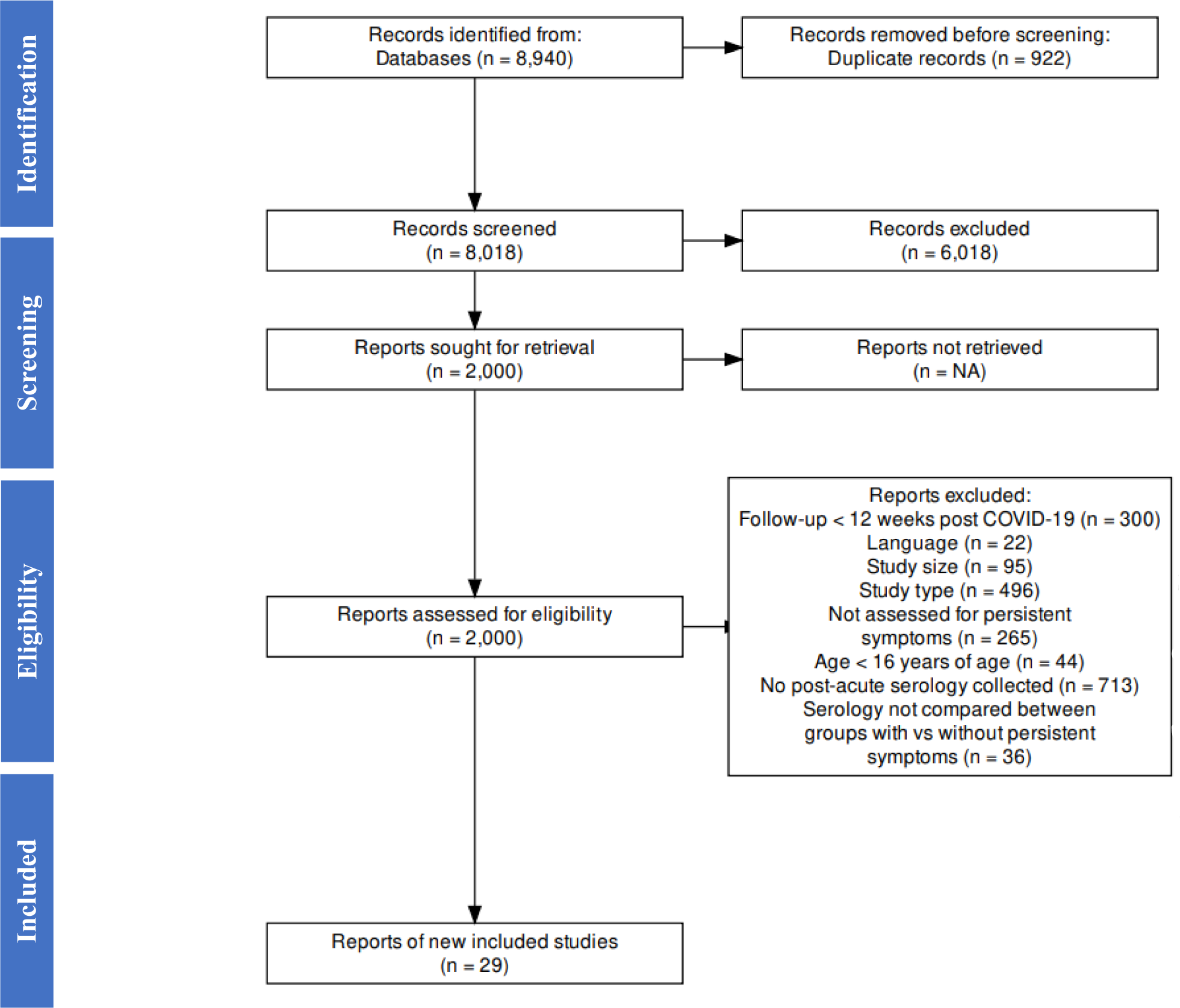
PRISMA diagram.

**Table 1:**
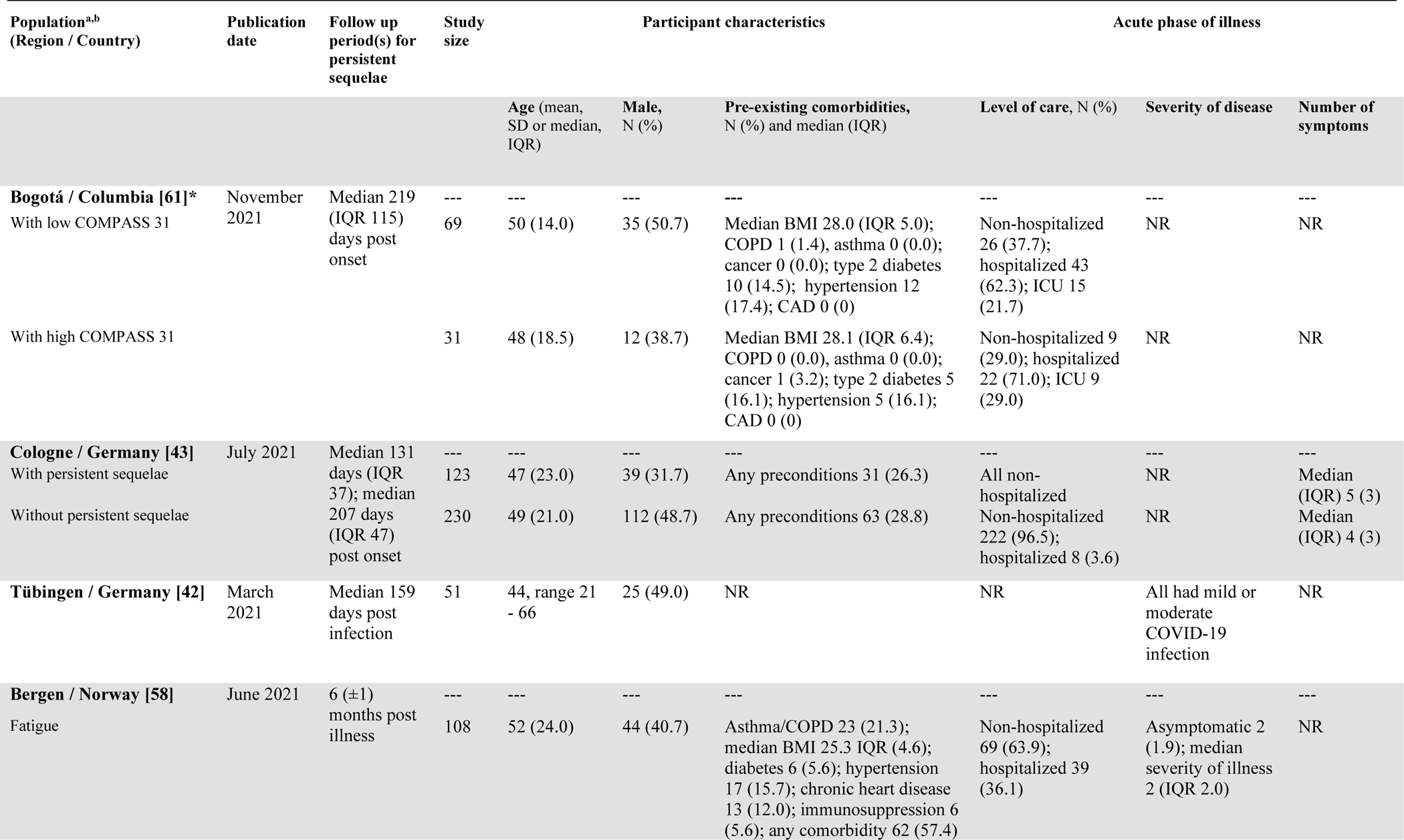
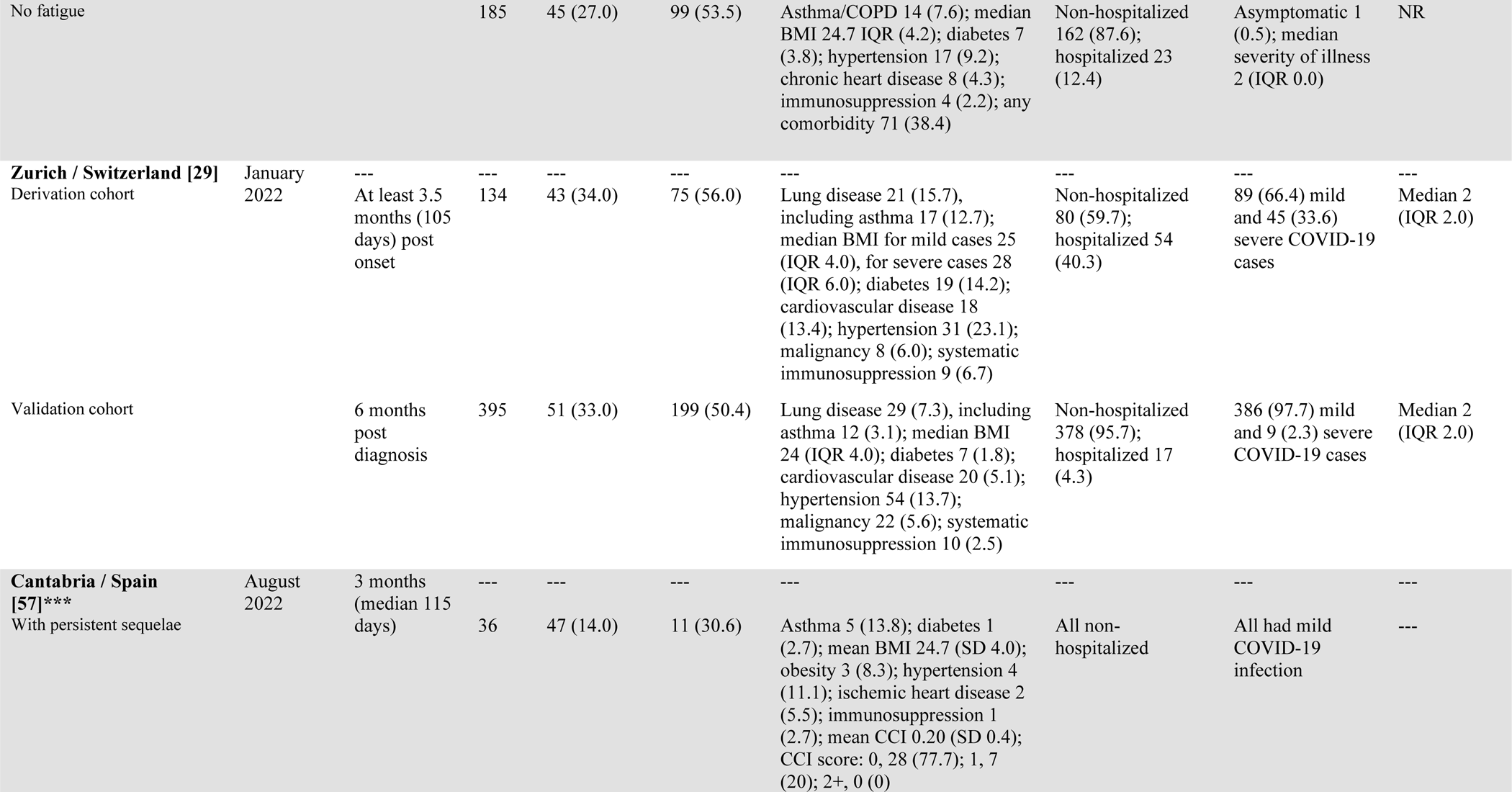
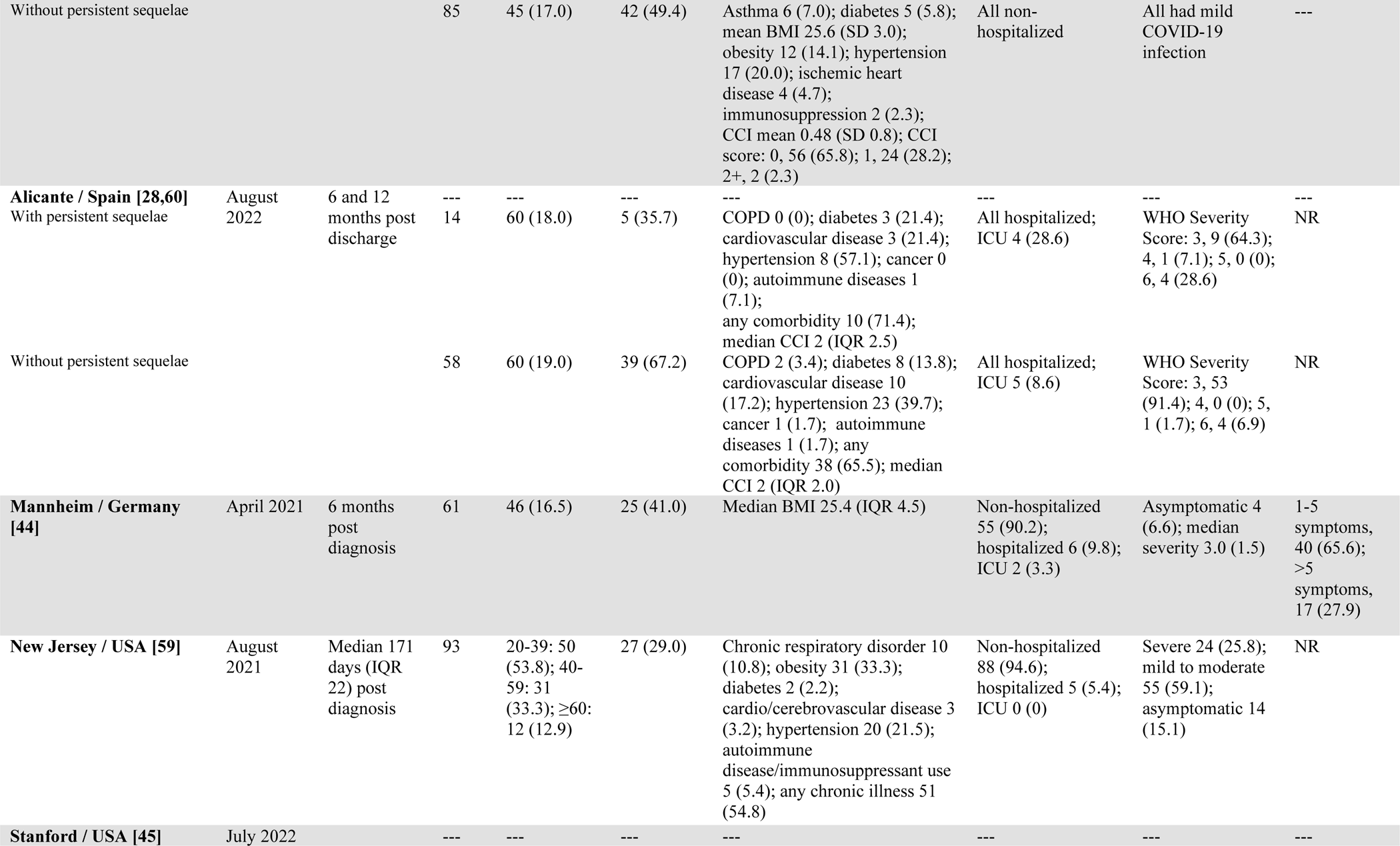
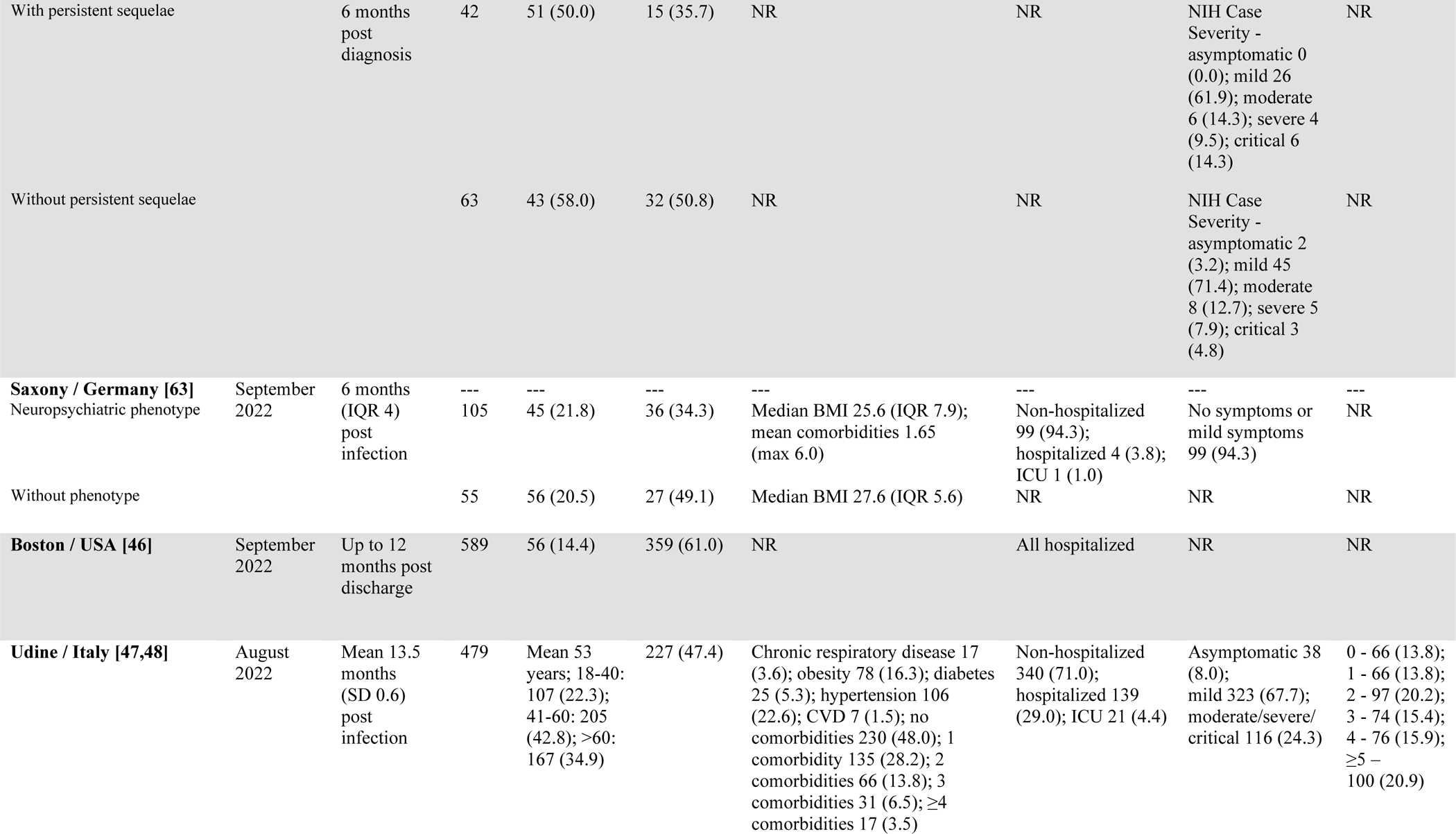
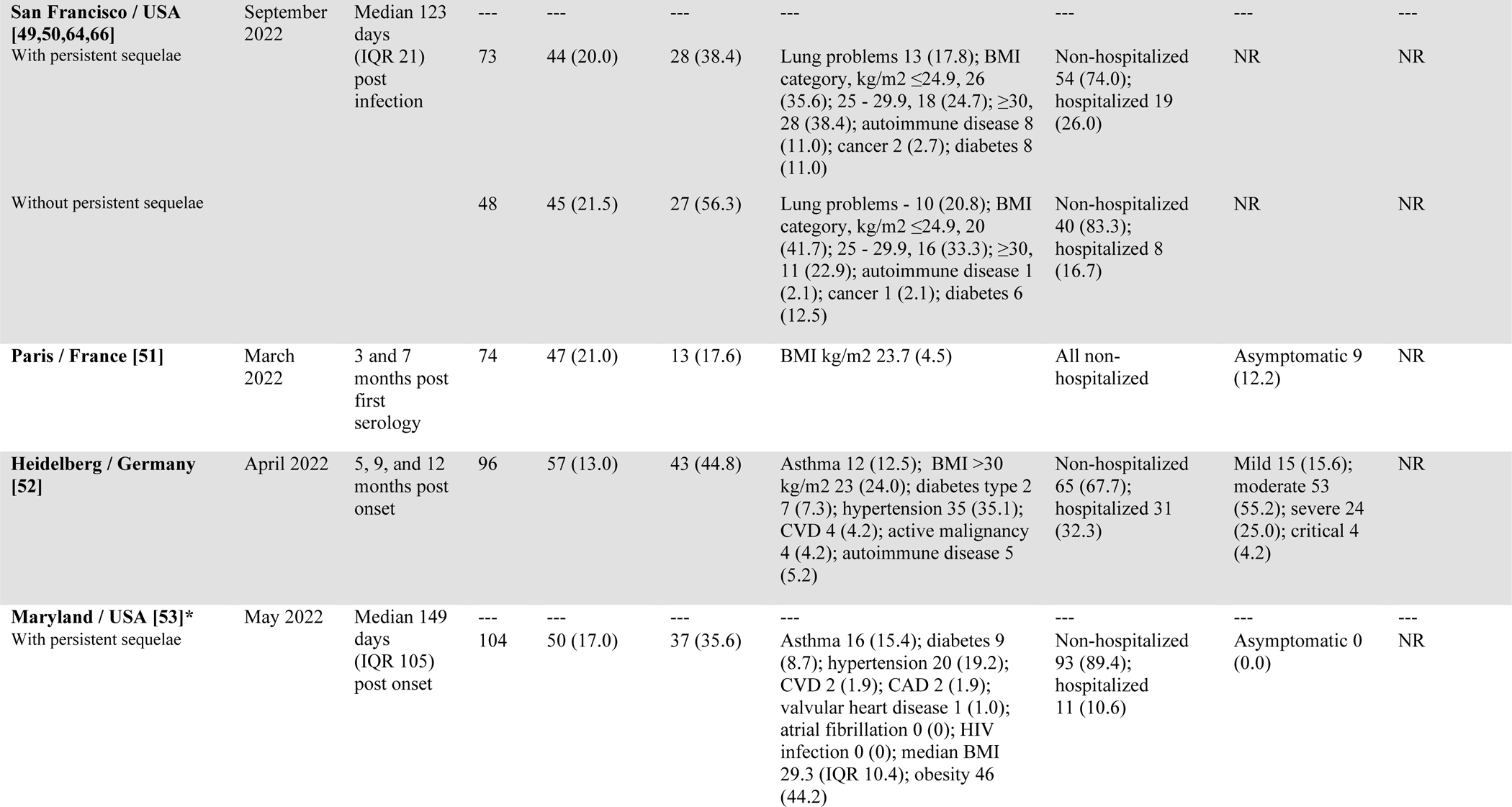
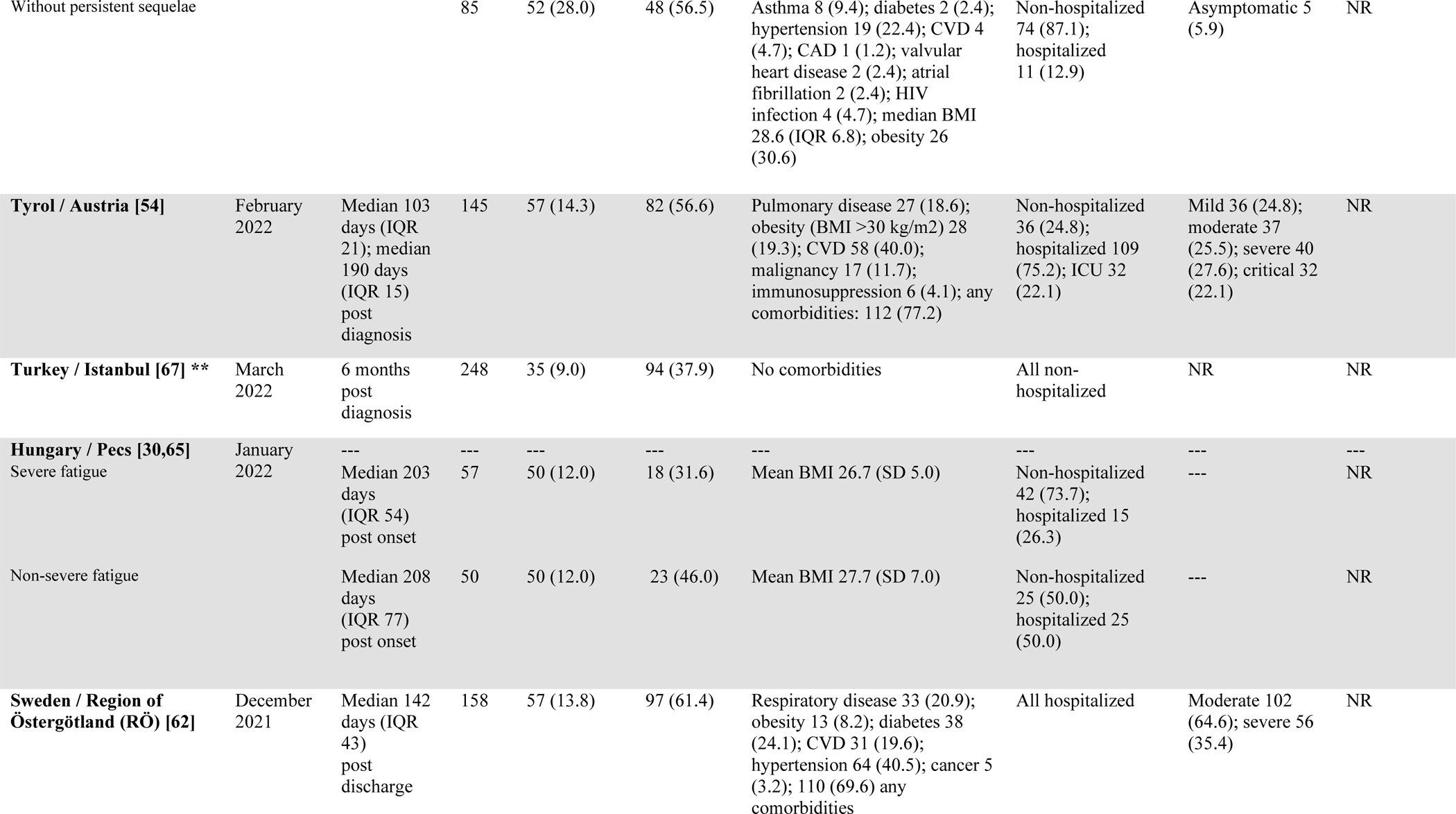
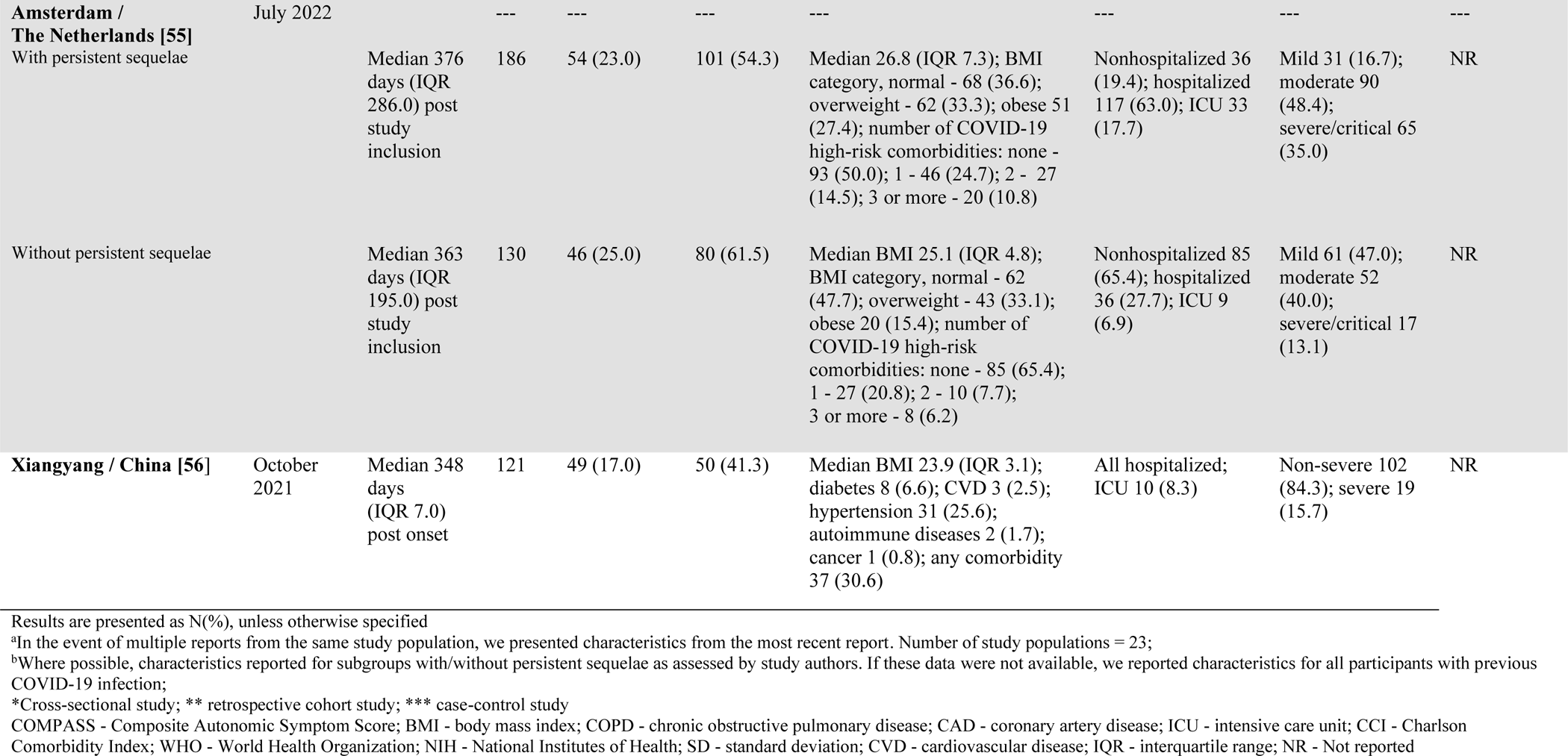
Characteristics of included studies (n=23 – 19 studies are of prospective cohort design; the design of the other four studies are indicted in footnotes)

### 5.4.2 Quality assessment

**Tables 2 – 4** display the quality grading of studies according to the NOS. Notably, most prospective cohort studies did not describe efforts to assess the outcome (persistent symptoms) prior to COVID-19 infection (n=15), and follow-up rate was often < 80% or not stated (n=14). Also, only 14 studies controlled for severity or LOC required during acute illness, while 17 studies assessed for other potential confounders (e.g., age, sex, pre-existing comorbidities), when assessing relationships between serological markers and PCC.

**Table 2:**
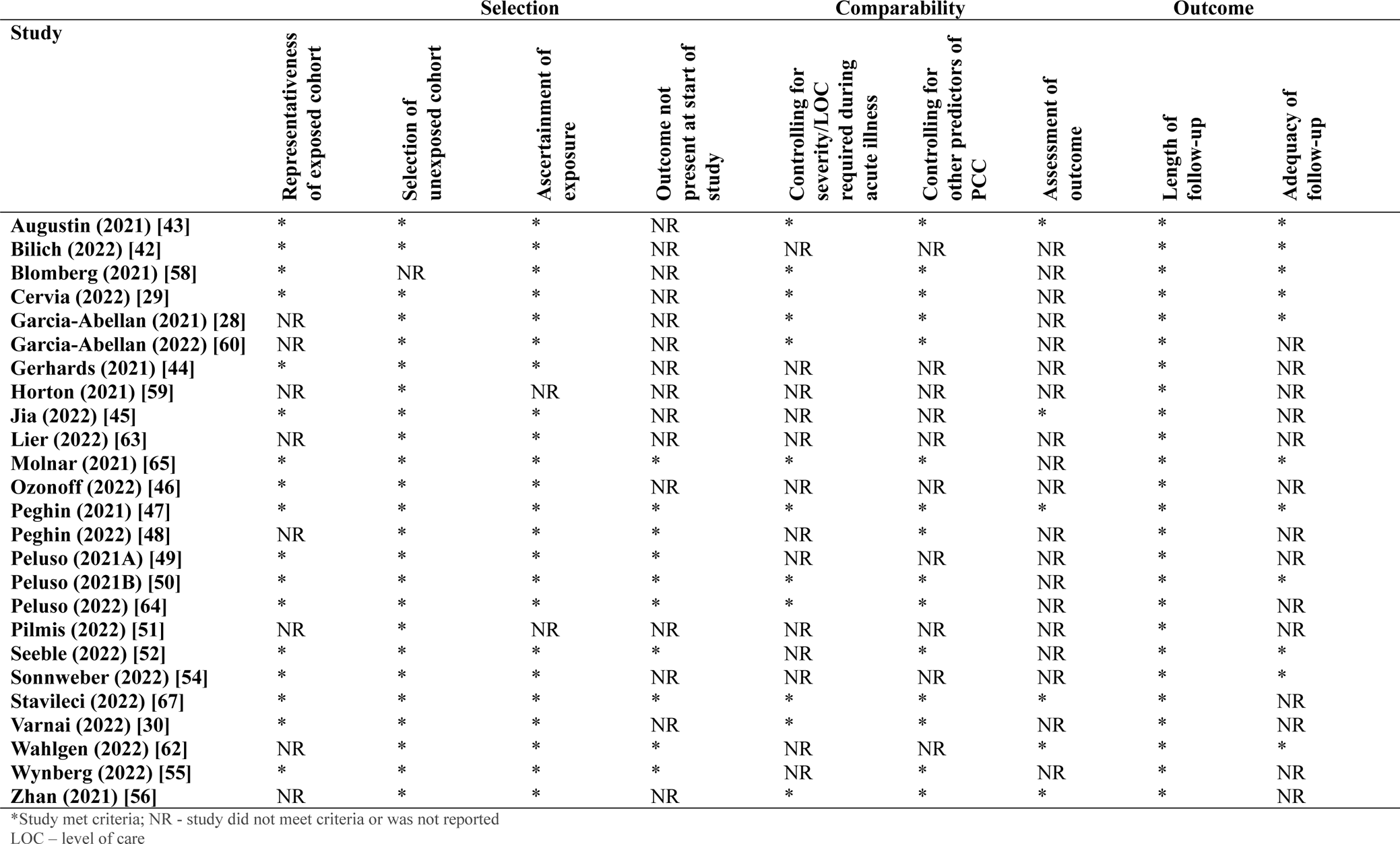
Quality assessment using the Newcastle-Ottawa Scale for cohort studies.

**Table 3:**
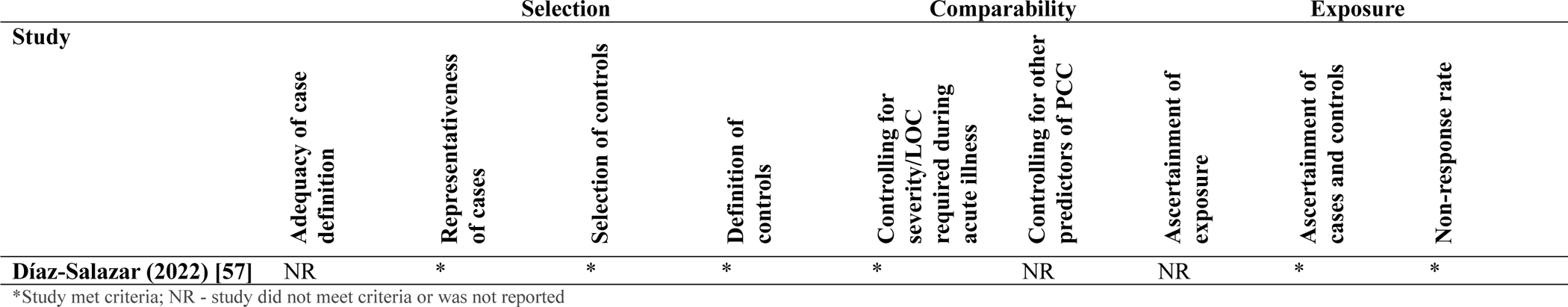
Quality assessment using the Newcastle-Ottawa Scale for case-control studies.

**Table 4:** Quality assessment using the Newcastle-Ottawa Scale for cross-sectional studies.

| Study | Selection |  |  | Comparability |  | Exposure |  |
| --- | --- | --- | --- | --- | --- | --- | --- |
|  | Representativeness of sample | Comparability between respondents and non-respondents | Ascertainment of exposure | Controlling for severity/LOC required during acute illness | Controlling for other predictors of PCC | Assessment of outcome | Statistical test |
| **Anaya (2021) [61]** | * | * | * | NR | NR | NR | * |
| **Durstenfeld (2022) [66]** | * | * | * | * | * | NR | * |
| **Sneller (2022) [53]** | * | * | * | NR | * | NR | NR |

\*Study met criteria; NR - study did not meet criteria or was not reported

### Persistent sequelae – definitions and subgroups for which serological comparisons available

#### Any symptoms vs no symptoms post COVID-19 onset

Studies used different strategies to define groups with and without persistent symptoms. Most commonly, studies compared findings among subgroups with any symptoms vs no symptoms following acute COVID-19 [29,42–44, 46–56,64]. Most study populations were assessed for PCC between 3 to < 6 months (n=8) or 6 to < 9 months (n=5) post COVID-19. Remaining populations were assessed for symptoms ≥12 months (n=4), or between three- and 12-months post COVID-19 (n=1). Some studies assessed PCC at multiple timepoints (**Table 1**).

#### Symptom duration and severity

Other studies reported findings based on symptom longevity and intensity of symptoms. A study on a working-age cohort [59] reported average antibody levels over time vs days post COVID-19 positive. Another study [45] assessed the association of antibody levels with time to sustained resolution for at least one month among a mixed population. Garcia-Abellan and colleagues [60] administered the COVID-19 symptoms questionnaire (CSQ), asking participants to self-report intensity of symptoms. Participants were classified as symptomatic if their score for any symptoms was in the top quartile of group scores.

#### PCC subtypes and clusters of PCC symptoms

Of nine studies to report on the presence or absence of specific symptoms/clusters, two [61,62] assessed for autonomic dysfunction, two [63,64] assessed for neurocognitive deficits, one assessed for sensorimotor impairments [62], three [30,58,65] assessed for fatigue, and two [66,67] assessed for cardiopulmonary symptoms.

### Serological results – trends by antibody type and target antigen

Serological results are summarized in **Tables S1 – S3** (by LOC required during acute illness), and **Tables S4 – S6** (by time interval (months) between COVID-19 infection and serological sampling). Below, we describe findings by antibody/target antigen, and discuss inter-study disparities which may have influenced results.

#### IgG response to SARS-CoV-2 nucleocapsid protein (**Tables S1** and **S4**)

Of 10 studies that assessed anti-N IgG response, only one controlled for acute disease severity/LOC [56]. Six studies (n=726 participants, **Figures S1a** and **S1b**) reported no difference in results between those with and without persistent symptoms post COVID-19, which were on mixed (n=2), hospitalized (n=2), and non-hospitalized (n=2) populations (**Figure 2a**). Three studies on mixed populations reported a decrease (n = 3 studies, 212 participants) of anti-N IgG among people with persistent symptoms, as compared to those without. **Figure 3a** displays the trend in anti-N IgG by time interval (months) between COVID-19 infection and serological sampling.

**Figure 2.**
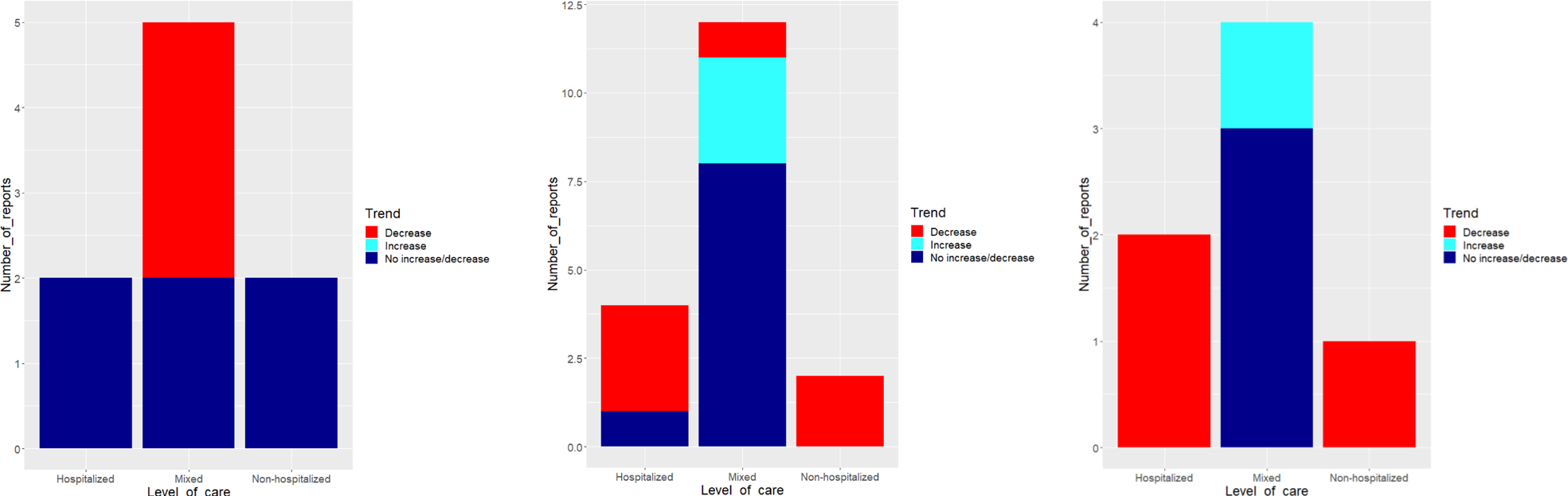
(a-c): Trends in serological response among groups with persistent symptoms as compared to groups without persistent symptoms, by level of care (LOC) requirements during acute illness. **a: Trend in anti-N IgG by level of care (LOC) requirements during acute illness** - number of studies to report any decrease, any increase, or no increase/decrease in anti-N IgG response among people with persistent symptom(s), as compared to people without persistent symptom(s). Of nine studies to assess anti-N IgG, two had hospitalized populations and two had non-hospitalized populations, all of which reported no increase/decrease. Of five studies with mixed populations to assess anti-N IgG, three studies reported ≥1 decrease and two studies reported no increase/decrease. One study [42] did not specify LOC and was hence excluded. **b: Trend in anti-Spike IgG by level of care (LOC) requirements during acute illness** - number of studies to report any decrease, any increase, or no increase/decrease in partial or full anti-Spike IgG response among people with persistent symptom(s), as compared to people without persistent symptom(s). Of 19 studies to assess full or partial anti-Spike IgG, four had hospitalized populations, of which one reported no increase/decrease, and three reported ≥1 decrease. Two studies had non-hospitalized populations, both of which reported ≥1 decrease. Finally, 12 studies had mixed populations, of which three reported ≥1 increase, one reported ≥1 decrease, and eight reported no increase/decrease. One study [42] did not specify LOC and was hence excluded. **c: Trend in neutralizing response by level of care (LOC) requirements during acute illness** - number of studies to report any decrease, any increase, or no increase/decrease in neutralizing response among people with persistent symptom(s), as compared to people without persistent symptom(s). Of seven studies to assess neutralizing response, two had hospitalized populations and one had a non-hospitalized population, all of which reported ≥1 decrease. The remaining four studies had mixed populations, one of which reported ≥1 increase with the remainder reporting no increase/decrease.

**Figure 3.**
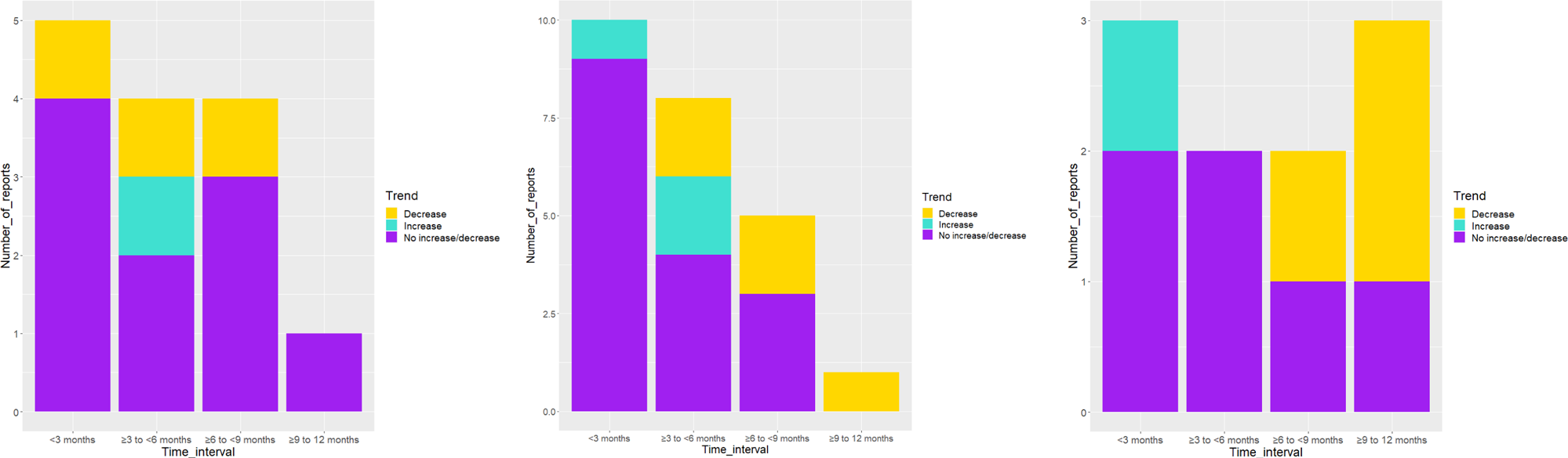
(a-c): Trends in serological response among groups with persistent symptoms as compared to groups without persistent symptoms, time interval (months) between COVID-19 infection and serological sampling. **a: Trend in anti-N IgG by time interval (months) between COVID-19 infection and serological sampling** - number of studies to report any decrease, any increase, or no increase/decrease in anti-N IgG response among people with persistent symptom(s), as compared to people without persistent symptom(s). Five studies assessed anti-N IgG <3 months post COVID-19, of which four reported no increase/decrease, and one reported ≥1 decrease. Four studies assessed anti-N IgG ≥3 months to <6 months post COVID-19, of which two reported no increase/decrease, one reported ≥1 increase, and one reported ≥1 decrease. Four studies assessed anti-N IgG ≥6 months to <9 months post COVID-19, of which three reported no increase/decrease, and one reported ≥1 decrease. Finally, one study assessed anti-N IgG ≥9 months to 12 months post COVID-19 and reported no increase/decrease. In studies with multiple timepoints of assessment, all results reported ≥12 weeks were included. **b: Trend in anti-Spike IgG by time interval (months) between COVID-19 infection and serological sampling** - number of studies to report any decrease, any increase, or no increase/decrease in partial or full anti-Spike IgG response among people with persistent symptom(s), as compared to people without persistent symptom(s). Ten studies assessed serology <3 months post COVID-19, of which one reported ≥1 increase and nine reported no increase/decrease. Eight studies assessed serology ≥3 months to <6 months post COVID-19, of which two reported ≥1 increase, two reported ≥1 decrease, and four reported no increase/decrease. Five studies assessed serology ≥6 months to <9 months post COVID-19, of which two reported ≥1 decrease, and three reported no increase/decrease. One study assessed serology ≥9 months up to 12 months post COVID-19, which reported ≥1 decrease. In studies with multiple timepoints of assessment, all results reported ≥12 weeks were included. **c: Trend in neutralizing response by time interval (months) between COVID-19 infection and serological sampling** - number of studies to report any decrease, any increase, or no increase/decrease in neutralizing response among people with persistent symptom(s), as compared to people without persistent symptom(s). Three studies assessed neutralizing response <3 months post COVID-19, of which two reported no increase/decrease, and one reported ≥1 increase. Two studies assessed neutralizing response ≥3 months to <6 months post COVID-19, of which both reported no increase/decrease. Two studies assessed neutralizing response ≥6 months to <9 months post COVID-19, of which one reported no increase/decrease increase/decrease and one reported ≥1 decrease. Finally, three studies assessed neutralizing response ≥9 months to 12 months post COVID-19, of which one reported no increase/decrease, and two reported ≥1 decrease. In studies with multiple timepoints of assessment, all results reported ≥12 weeks were included.

#### IgG response to SARS-CoV-2 Spike protein, S1/S2 subunits, and RBD (**Tables S2** and **S5**)

Of 19 studies to assess full or partial anti-Spike IgG response, 10 studies (n=1470 participants, **Figures S1a** and **S1b**) reported no increase/decrease between people with vs without persisting symptoms. These studies had mixed or hospitalized populations (**Figure 2b**), and most (n = 10) sampled serology < 3 months post-infection (**Figure 3b**). Three studies (459 participants) found increased titres among people with persistent symptoms as compared to those without symptoms, all of which had mixed study populations (**Figure 2b**). Six studies (851 participants) reported ≥ 1 decrease in serological results among people with persistent symptoms as compared to those without symptoms, and assessed mixed (n=1), hospitalized (n=3), and non-hospitalized (n=2) populations.

#### Neutralizing antibodies (**Tables S3** and **S6**)

Of seven studies to assess neutralizing response, three studies (n=477 participants) reported no difference. Three studies (n=353 participants) reported ≥ 1 decrease. One study (n=293 participants) found microneutralizing titres assessed two months post-infection to be positively associated with fatigue score, controlling for severity of acute illness and other covariates. **Figure 3c** displays the trend in neutralizing titres by time interval (months) between COVID-19 infection and serological sampling.

#### IgM and IgA response

Few studies (n=2; 629 participants) reported on IgM and/or IgA response to SARS-CoV-2 antigens, which has been noted previously [68], with respect to PCC status. Anaya et al. [61] compared median (U/mL)/ % of patients anti-RBD IgG, IgA, and IgM between participants with low COMPASS 31 (Composite Autonomic Symptom Score) scores (Cluster 1) as compared to participants with high COMPASS 31 scores (Cluster 2) and found results to be non-significant (p=0.24). Cervia et al. [29] found the log odds of an interaction term between IgM and IgG3 to be negatively associated with persisting symptoms (−2.13, 95% CI −4.45, −0.29) among a mixed population, accounting for covariates.

### Vaccination status

Of 13 studies reporting vaccination status, seven reported all participants to be non-vaccinated, and six reported vaccination prior to study recruitment and/or during the study. Of 16 studies to not report vaccination status, most (n=14) recruited participants infected prior to mass-vaccination. Of the six studies to report any vaccination, all participants in five studies were infected prior to mass-vaccination and < 5% of participants in the sixth study completed two vaccine doses prior to baseline visit. Two studies [30,48] compared results for vaccinated and non-vaccinated subgroups.

### COVID-19 variant

Only one study reported the COVID-19 strain(s) that infected study participants. Where not specified, we inferred strains to be those which prevailed in the host country of the study during infection or recruitment dates [39]. If these dates were not indicated by the study, we identified the dominant strains to have preceded data collection post-infection. Through this process, we determined that all studies recruited participants to have been infected when wild-type or alpha strains prevailed. Two studies [63,65] may also have recruited participants who were infected when the delta variant was the dominant strain.

## Discussion

As part of the global research response to the pandemic, many studies collected data on humoral response following COVID-19 infection. A subset of these studies also examined for persisting symptoms. Such endeavours demand extensive commitments of time and effort from multidisciplinary research groups, and necessitate substantial funds for study design, implementation, and maintenance. Notably, neutralization assays can be especially costly and labour-intensive [71,72].

A multitude of factors can influence PCC and post-infection serological trends. Controlling for potential confounders is a critical prerequisite to establishing the magnitude and direction of relationships between serological markers and PCC [3,8,17,31]. Given the considerable clinical and processing throughput required of eligible studies, large sample sizes with blood draws at multiple timepoints may not be feasible. Pooling of inter-study findings would enable more robust exploration of multiple clinical and serological predictors among varying populations.

For these reasons, we performed a rapid review of serological markers which may be associated with PCC, and summarized variations which hampered comparability of inter-study findings. Given substantial heterogeneity in participant characteristics, study procedures, and serological parameters, we were not able to pool results. Upon reviewing overall trends for anti-N IgG, full or partial anti-Spike IgG, and neutralizing response, we inferred the following:

1. Results suggest no difference in anti-N IgG by PCC status. Studies which reported any increase/decrease were studies with mixed populations that did not account for initial disease severity or LOC. Hence, differences in anti-N IgG response may have been driven by response in the initial phase of illness.
2. Studies on populations with varying LOC requirements and time intervals of serological sampling ≥3 months post-infection (**Figures 3b** and **4b**) reported ≥1 decrease, when comparing full or partial anti-Spike response among individuals with persisting symptoms to response among individuals without. However, PCC definitions and the analyses and reporting of results were highly variable. Therefore, we can neither refute nor confirm evidence of differences by PCC status.
3. Seven studies assessed neutralizing response. Results were highly variable. Of four studies to report any difference in findings by PCC status, only one study compared results between groups of people with any symptoms vs no symptoms. The remaining three studies assessed for differences by PCC phenotype, severity, or the presence of specific symptoms (e.g., fatigue).
4. A small subset of studies examined specific symptom(s) or symptom clusters. Further investigation of these findings may elucidate new insights otherwise obscured by use of a blanket definition of PCC. For example, the one study to compare humoral response among groups with and without dyspnea, chest pain, or palpitations reported increased odds of symptoms per doubling of anti-RBD levels, accounting for covariates [66]. Studies to assess fatigue found decreased anti-N IgG among those with severe fatigue as compared to those with non-severe fatigue, and increased risk of fatigue status given higher microneutralizing titres. Finally, the one study to assess neutralizing response among groups with and without a neuropsychiatric phenotype reported decreased neutralizing antibodies among those with symptoms [63].

### Recommendations to improve the quality and comparability of evidence

Findings are largely inconclusive as the bulk of evidence failed to account for potential confounders and there are substantial inter-study inconsistencies. We propose the following recommendations to improve the quality and comparability of findings on post-infection serology and PCC:

#### Sharing of serological results collected at common timepoints post-infection, guided by knowledge of expected rates of seroconversion and decay

Serological sampling timepoints varied considerably, given no accepted standards [73,74]. Results may differ depending on months post-infection at which blood is collected for serological analysis [74–76]. This is especially true if sampling timepoints vary between groups with and without persisting symptoms. We propose that the expected trajectory of immunoglobulins post COVID-19 infection warrants consideration when interpreting serological findings from different post-infection timepoints. Seroconversion for all antibody types occurs on average four to 14 days post-onset [76]. A systematic review of post-infection humoral response found IgG to be detected an average of 12 days post onset, to peak at 25 days, and to start to decline after two months [77]. Seronegative results are more likely prior to 14 days or after six months post-infection [69, 74, 75,76–79]. Target antigen and severity of acute disease may also influence rate of decay [69,74–76]. Multiple studies have found anti-N IgG response to decay more rapidly than response to Spike [74–76]. Furthermore, individuals who experience mild COVID-19 disease are less likely to develop detectable antibodies and more likely to exhibit delayed IgG seroconversion, as compared to those with more severe COVID-19 [80–82].

#### A consensus on analyses and reporting of serological results

To better enable harmonization of results from different assays, the WHO’s Expert Committee on Biological Standardization developed an International Standard and Reference Panel for SARS-CoV-2 antibodies [82]. Serological findings recalibrated on this standard are reported as binding antibody units (BAU/mL). However, some studies have found differences in recalibrated results derived from different assays [73,83]. Additionally, variable derivation of cut-offs and thresholds and units of quantitative results obscure understanding of findings. Studies that report strength of response using cut-offs (e.g., low, medium, or high titres) should delineate cut-offs as pre-specified or exploratory, and explain how they were derived [76–78]. Also, endeavours to assess serological decay may only state whether or not there was a difference in results over time: absolute values should be reported to improve transparency and comprehension of findings. Finally, given the importance of collaboration across multiple disciplines to advance knowledge on PCC, there is great need for clear communication and shared understanding around the meaning and limitations of findings [76–78,84].

#### More reports on specific PCC symptoms and symptom clusters

Knowledge of PCC continues to evolve, as do the definitions for this condition and subtypes based on varying severity or character of symptoms [1–4,6]. The exploration of PCC subtypes is an important and emerging topic, with potential to advance our understanding of pathophysiological mechanisms and markers, and better enable health systems to identify and address key care needs [6,84]. However, there continues to be poor consensus on what these subtypes are, and how clinical characteristics and COVID-19 variants may influence the manifestation and severity of different symptom patterns [6,33,85,86]. More reports on subtypes and potential biomarkers may yield new findings which illuminate PCC etiology, detection, and treatment.

#### Risk of bias – recommendations to improve the quality of evidence

Common factors threatening study quality included failures to describe efforts to confirm that SARS-CoV-2 infection preceded the outcome, and to control for potential confounders. We identified acute severity of illness as the most important confounder to consider, given that substantial evidence has highlighted this to be a major driver of serological response [69,70,73,76,81,87], and many studies have found more severe illness early on to be predictive of PCC onset and trajectory [15,16,20,88].

Some studies also restricted serological follow-up to seropositive cases. This strategy may have biased results towards the null. Results are more likely to have been influenced if seropositivity was determined prior to the generation of detectable antibodies post-infection, or after antibodies and sensitivity begin to diminish, depending on assay and severity of acute illness [74].

### Strengths and limitations

Key strengths of this review include the large volume of reports assessed for eligibility, and careful consideration and thorough description of a wide array of factors which limit inter-study comparability. Also, we reported findings among different PCC subtypes, currently an important and growing area of research interest [85]. However, several limitations warrant consideration. First, we noted restricted variation in terms of COVID-19 strain and vaccination status. The majority of participants from all studies were infected by wildtype/alpha strains, and vaccine naive at time of infection. Therefore, there was limited opportunity to explore the effects of hybrid immunity and different variants of concern on findings. Second, the literature on PCC and COVID-19 immune response continues to evolve; evidence published after our search date in October 2022 may yield different findings. Third, given variations in serological response and PCC presentation among children, we chose to focus this review on adult COVID-19 survivors [89,90]. Therefore, our results are not generalizable to younger age groups. Fourth, the synthesis only focused on IgG response to SARS-CoV-2 antigens and measures of neutralizing efficiency. Other potential biomarkers were not explored. Fifth, we acknowledge the risk of survivor bias, especially among studies on hospitalized populations. Ozonoff et al. [46] found that patients who died during acute illness had lower antibody titres than survivors, many of whom went on to be assessed ≥12 weeks post-infection. Sixth, this review focused on self-reported symptoms and severity. While this approach integrates the patient perspective, there is risk of reporting bias. Finally, we did not assess for effects from COVID-19 re-infections.

## Conclusion

Examination of PCC onset and phenotype as functions of serological predictors, accounting for clinical covariates, may yield emergent insights and advance understanding of PCC etiology, detection, and treatment. As the assessment of COVID-19 humoral response is not a standard practice in healthcare settings, serological results by PCC status have been made available through international research efforts. However, given poor consensus on standards of clinical and serological collection, analysis, and reporting, there are substantial inter-study inconsistencies. Uniform efforts to harmonize reporting of serological results and control for acute disease severity or level of care requirements would improve the quality, comparability, and comprehension of findings. There is also continued need for reports on PCC subtypes, an important and evolving topic with potential to advance understanding of pathophysiological mechanisms and markers, and better enable health systems to identify and address key care needs. Finally, future reviews of ongoing studies will facilitate more detailed analyses of the effects of SARS-CoV-2 vaccination and variants on findings.

## Acknowledgements

The search strategy was developed with assistance from the Canadian Health Library.

## Author contributions

EC and JL drafted the manuscript. EP assisted screening, data extraction, and risk of bias assessment. CG, SH, and MAL provided expertise on epidemiological and serological content. All authors critically reviewed and approved the final manuscript.

## Data availability

All relevant data are within the manuscript. No additional source data are required.

