## Supplementary Materials for "Serological markers and Post COVID-19 Condition (PCC) – A rapid review of the evidence"

| 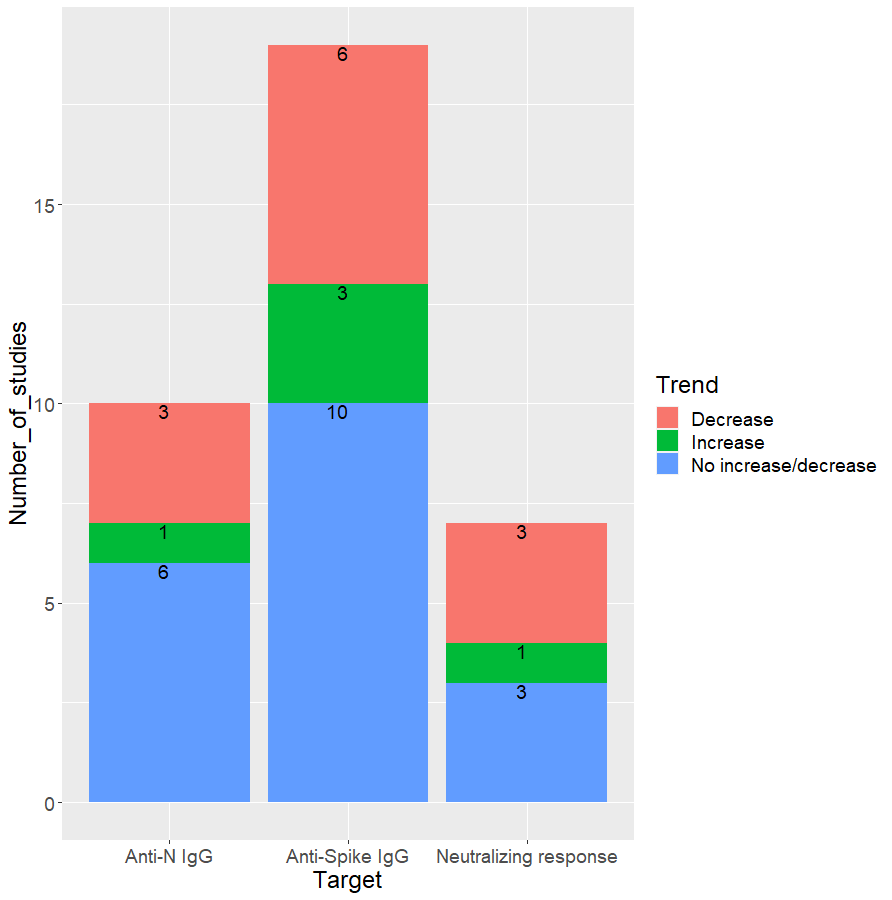 | 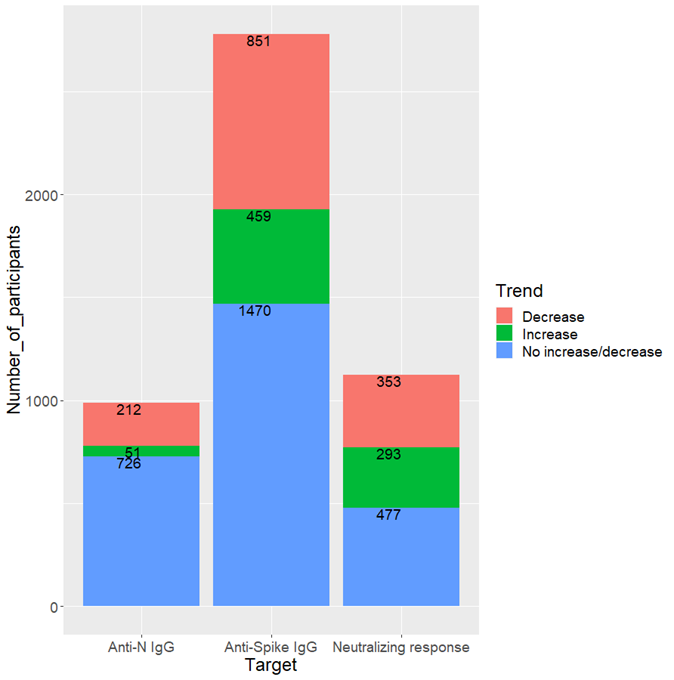 |
| --- | --- |
| **Figure S1a:** Number of studies to report any decrease, any increase, or no increase/decrease in serological findings among individuals with persistent symptoms, as compared to individuals without persistent symptoms. | **Figure S1b:** Aggregate number of unique participants in studies to report any decrease, any increase, or no increase/decrease in serological findings among individuals with persistent symptoms, as compared to individuals without persistent symptoms. In cases where there were multiple reports per study population, we reported the number of participants in the initial report. |

| **Table S1: Anti-N IgG comparisons among groups with and without persistent symptoms** **≥12 weeks post COVID-19, stratified by level of care requirements during the acute phase of illness** | | | | | | | | | |
| --- | --- | --- | --- | --- | --- | --- | --- | --- | --- |
| **Study population** | | **Measure / unit** | **Timepoints assessed** | | **Comparisons** | | | | **Overall trend^a^** |
|  | |  | **Serology** | **Post-acute** | **Group** | **Participants**  **assessed (N)** | **Results** | **Variables adjusted** |  |
| **Hospitalized (studies = 2; participants = 237)** | | | | | | | | | |
| García-Abellán, Alicante / Spain [60] | | N (%) seropositive | 1 month | 6 months | Highest CSQ scores after discharge | 28 | 17 (68.0) | NA | No increase  / decrease |
| García-Abellán, Alicante / Spain [60] | | N (%) seropositive | 1 month | 6 months | Other participants | 88 | 59 (74.0) | NA | No increase  / decrease |
| García-Abellán, Alicante / Spain [60] | | N (%) seropositive | 2 months | 6 months | Highest CSQ scores after discharge | 28 | 17 (68.0) | NA | No increase  / decrease |
| García-Abellán, Alicante / Spain [60] | | N (%) seropositive | 2 months | 6 months | Other participants | 88 | 61 (72.6) | NA | No increase / decrease |
| García-Abellán, Alicante / Spain [60] | | N (%) seropositive | 6 months | 6 months | Highest CSQ scores after discharge | 28 | 19 (73.1) | NA | No increase / decrease |
| García-Abellán, Alicante / Spain [60] | | N (%) seropositive | 6 months | 6 months | Other participants | 88 | 61 (71.8) | NA | No increase / decrease |
| García-Abellán, Alicante / Spain [60] | | Peak, serum  concentration - S/CO median (IQR) | 1, 2, 6 months | 6 months | Highest CSQ scores after discharge | 28 | 3.8 (2.9) | NA | No increase / decrease |
| García-Abellán, Alicante / Spain [60] | | Peak, serum  concentration - S/CO median (IQR) | 1, 2, 6 months | 6 months | Other participants | 88 | 4.1 (2.2) | NA | No increase / decrease |
| García-Abellán, Alicante / Spain [60] | | Trough, serum  concentration - S/CO median (IQR) | 1, 2, 6 months | 6 months | Highest CSQ scores after discharge | 28 | 1.7 (1.3) | NA | No increase / decrease |
| García-Abellán, Alicante / Spain [60] | | Trough, serum  concentration - S/CO median (IQR) | 1, 2, 6 months | 6 months | Other participants | 88 | 2.2 (1.2) | NA | No increase / decrease |
| Zhan, Xiangyang / China [56] | | Beta (95% CI) | 10-12 months | 10-12 months | Any persistent symptoms vs no persistent symptoms | 121 | -0.16  (-0.34, 0.01) | Time from discharge, length of stay, age, sex, severe disease, glucocorticoids, interferons | No increase / decrease |
| **Non-hospitalized (studies = 2; participants = 234)** | | | | | | | | | |
| Lier, Saxony / Germany [63] | | Median (IQR), (S/CO) | 6-9 months | 6 months | Neuropsychiatric phenotype | 105 | 1.4 (1.2) | NA | No increase / decrease |
| Lier, Saxony / Germany [63] | | Median (IQR), (S/CO) | 6-9 months | 6 months | Without phenotype | 55 | 1.4 (1.3) | NA | No increase / decrease |
| Pilmis, Paris / France [51] | | Inter-group comparisons using the Mann-Whitney test | 1, 3, 8 months | 3 months | Any persistent symptoms vs no persistent symptoms | 74 | Not specified | NA | No increase / decrease |
| **Mixed (studies = 5; participants = 467)** | | | | | | | | | |
| Jia, Stanford / California, USA [45] | | N (%), (High > 149,452 AU/mL) | In the first month of illness | 6 months | Probability of reporting persistent symptoms | NR | 55.0 % | NA | No increase / decrease |
| Jia, Stanford / California, USA [45] | | N (%), (Low <=149,452 AU/ml) | In the first month of illness | 6 months | Probability of reporting persistent symptoms | NR | 36.0 % | NA | No increase / decrease |
| Jia, Stanford / California, USA [45] | | Hazard Ratio (95% CI) | In the first month of illness | 6 months | Percentage of participants with resolved symptoms vs days post COVID-19 diagnosis | 105 | 3.5  (1.0, 11.6) | NA | Decrease |
| Molnar, Hungary / Pecs [65] | | Median (IQR), (U/mL) | > 3 months | > 3 months | Severe fatigue | 63 | 24.8 (50.0) | NA | Decrease |
| Molnar, Hungary / Pecs [65] | | Median (IQR), (U/mL) | > 3 months | > 3 months | Less severe fatigue | 38 | 90.0 (60.0) | NA | Decrease |
| Sneller, Maryland / USA [53] | | N (%) | 5-6 months | 5-6 months | Any persistent symptoms | 104 | 90 (86.5) | NA | No increase / decrease |
| Sneller, Maryland / USA [53] | | N (%) | 5-6 months | 5-6 months | No persistent symptoms | 85 | 69 (81.2) | NA | No increase / decrease |
| Varnai, Hungary / Pecs [30] | | Median (IQR), (U/mL) | 6-7 months | 6-7 months | Severe fatigue | 57 | 27.0 (75.0) | NA | Decrease |
| Varnai, Hungary / Pecs [30] | | Median (IQR), (U/mL) | 6-7 months | 6-7 months | Non-severe fatigue | 50 | 98.0 (123.0) | NA | Decrease |
| Peluso, San Francisco / California [49] | | Differences in median anti-N (RU) at T1 and T4 | 1-2 months to 4-5 months | 4 months | Any persistent symptoms vs no persistent symptoms | 66 | Not specified | NA | No increase / decrease |
| ^a^Trend as reported by study authors  CSQ - COVID-19 symptoms questionnaire; IQR - interquartile range; RU - relative light units; S/CO - signal to cut-off ratio; NA - Not applicable; NR - Not reported; T1 - Visit 1; T4 - Visit 4; OR - odds ratio; CI - confidence interval | | | | | | | | | |

| **Table S2: Anti-Spike IgG comparisons (Spike protein, subunits, or RBD) among groups with and without persistent symptoms ≥12 weeks post COVID-19, stratified by level of care requirements during the acute phase of illness** | | | | | | | | |
| --- | --- | --- | --- | --- | --- | --- | --- | --- |
| **Study cohort** | **Measure / unit** | **Timepoints assessed** | | **Comparisons** | | | | **Overall trend^a^** |
|  |  | **Serology** | **Post-acute** | **Group** | **Participants assessed (N)** | **Results** | **Variables adjusted** |  |
| **Hospitalized (studies = 4; participants = 826)** | | | | | | | | |
| García-Abellán, Alicante / Spain [60] | N (%) IgG-Spike seropositive | 1 month | 6 months | Highest CSQ scores after discharge | 28 | 17 (68.0) | NA | No increase / decrease |
| García-Abellán, Alicante / Spain [60] | N (%) IgG-Spike seropositive | 1 month | 6 months | Other participants | 88 | 58 (72.5) | NA | No increase / decrease |
| García-Abellán, Alicante / Spain [60] | N (%) IgG-Spike seropositive | 2 months | 6 months | Highest CSQ scores after discharge | 28 | 17 (68.0) | NA | No increase / decrease |
| García-Abellán, Alicante / Spain [60] | N (%) IgG-Spike seropositive | 2 months | 6 months | Other participants | 88 | 61 (72.6) | NA | No increase / decrease |
| García-Abellán, Alicante / Spain [60] | N (%) IgG-Spike seropositive | 6 months | 6 months | Highest CSQ scores after discharge | 28 | 19 (73.1) | NA | No increase / decrease |
| García-Abellán, Alicante / Spain [60] | N (%) IgG-Spike seropositive | 6 months | 6 months | Other participants | 88 | 62 (72.9) | NA | No increase / decrease |
| García-Abellán, Alicante / Spain [60] | Peak IgG-Spike, serum concentration - S/CO (IQR) | 1, 2, 6 months | 6 months | Highest CSQ scores after discharge | 28 | 5.1 (3.9) | NA | No increase / decrease |
| García-Abellán, Alicante / Spain [60] | Peak IgG-Spike, serum concentration - S/CO (IQR) | 1, 2, 6 months | 6 months | Other participants | 88 | 6.3 (4.6) | NA | No increase / decrease |
| García-Abellán, Alicante / Spain [60] | Trough IgG-Spike, serum concentration - S/CO (IQR) | 1, 2, 6 months | 6 months | Highest CSQ scores after discharge | 28 | 3.9 (28.0) | NA | No increase / decrease |
| García-Abellán, Alicante / Spain [60] | Trough IgG-Spike, serum concentration - S/CO (IQR) | 1, 2, 6 months | 6 months | Other participants | 88 | 3.7 (2.5) | NA | No increase / decrease |
| García-Abellán, Alicante / Spain [60] | Peak IgG-Spike S/CO values, OR (95% CI) | Up to 6 months | 6 months | Highest CSQ scores after discharge vs other participants | 116 | 0.9  (0.79, 0.99) | Age, sex, Charlson comorbidity index, WHO severity ordinal scale score, testing positive for SARS-CoV-2 RT-PCR at 1-month visit and tocilizumab use | Decrease |
| García-Abellán, Alicante / Spain [60] | Peak IgG-Spike S/CO values, OR (95% CI) | Up to 6 months | 6 months | Median or above median CSQ score after discharge vs other participants | 116 | 0.9  (0.78, 1.07) | Age, sex, Charlson comorbidity index, WHO severity ordinal scale score, testing positive for SARS-CoV-2 RT-PCR at 1-month visit and tocilizumab use | No increase / decrease |
| García-Abellán, Alicante / Spain [60] | Peak IgG-Spike S/CO values, OR (95% CI) | Up to 6 months | 6 months | Any symptom after discharge vs other participants | 116 | 1.0  (0.90, 1.20) | Age, sex, Charlson comorbidity index, WHO severity ordinal scale score, testing positive for SARS-CoV-2 RT-PCR at 1-month visit and tocilizumab use | No increase / decrease |
| García-Abellán, Alicante / Spain [28] | Median (IQR) anti-S1/S2 (AU/Ml) | 12 months | 6 and 12 months | Highest CSQ scores after discharge | 14 | 49.0 (80.0) | NA | No increase / decrease |
| García-Abellán, Alicante / Spain [28] | Median (IQR) anti-S1/S2 (AU/Ml) | 12 months | 6 and 12 months | Other participants | 58 | 96.3 (86.6) | NA | No increase / decrease |
| García-Abellán, Alicante / Spain [28] | Median (IQR) anti-Spike (S/CO) | 12 months | 6 and 12 months | Highest CSQ scores after discharge | 14 | 1.9 (3.3) | NA | No increase / decrease |
| García-Abellán, Alicante / Spain [28] | Median (IQR) anti-Spike (S/CO) | 12 months | 6 and 12 months | Other participants | 58 | 3.3 (2.7) | NA | No increase / decrease |
| García-Abellán, Alicante / Spain [28] | aHR (95% CI) anti-SI/S2, (AU/Ml) | 12 months | 6 and 12 months | Highest CSQ scores after discharge vs other participants | 72 | 0.1  (0.03, 0.65) | Sex and ICU stay | Decrease |
| Ozonoff,  Boston / USA [46] | Anti-RBD over time: trend lines representing the fit of a generalized additive model (GAM) | 1 month | 3, 6, 9, and/or 12 months | Any persistent symptoms vs no persistent symptoms | 589 | Not specified | NA | No increase / decrease |
| Zhan, Xiangyang / China [56] | Beta (95% CI), total anti-RBD | 10-12 months | 10-12 months | Any persistent symptoms vs no persistent symptoms | 121 | -0.35  (-0.57,  -0.13) | Time from discharge, length of stay, age, sex, severe disease, glucocorticoids, interferons | Decrease |
| Zhan, Xiangyang / China [56] | Beta (95% CI),  anti-RBD | 10-12 months | 10-12 months | Any persistent symptoms vs no persistent symptoms | 121 | -0.21 (-0.38,  -0.05) | Time from discharge, length of stay, age, sex, severe disease, glucocorticoids, interferons | Decrease |
| **Non-hospitalized (studies = 2; participants = 513)** | | | | | | | | |
| Augustin, Germany / Cologne [43] | S/CO-Ratio, median (IQR) anti-S1 | 1-2 months | 6-7 months | Any persistent symptoms | 123 | 3 (5.0) | NA | NR |
| Augustin, Germany / Cologne [43] | S/CO-Ratio, median (IQR) anti-S1 | 4-5 months | 6-7 months | Any persistent symptoms | 123 | 2 (3.0) | NA | NR |
| Augustin, Germany / Cologne [43] | S/CO-Ratio, median (IQR) anti-S1 | 6-7 months | 6-7 months | Any persistent symptoms | 123 | 2 (3.0) | NA | NR |
| Augustin, Germany / Cologne [43] | S/CO-Ratio, median (IQR) anti-S1 | 1-2 months | 6-7 months | No persistent symptoms | 230 | 4 (5.0) | NA | NR |
| Augustin, Germany / Cologne [43] | S/CO-Ratio, median (IQR) anti-S1 | 4-5 months | 6-7 months | No persistent symptoms | 230 | 3 (3.0) | NA | NR |
| Augustin, Germany / Cologne [43] | S/CO-Ratio, median (IQR) anti-S1 | 6-7 months | 6-7 months | No persistent symptoms | 230 | 2 (3.0) | NA | NR |
| Augustin, Germany / Cologne [43] | Anti-S1 low  (≤1.1 S/CO), N (%) | 6-7 months | 6-7 months | Any persistent symptoms | 123 | 21 (17.6) | NA | NR |
| Augustin, Germany / Cologne [43] | Anti-S1 low  (≤1.1 S/CO), N (%) | 6-7 months | 6-7 months | No persistent symptoms | 230 | 28 (12.8) | NA | NR |
| Augustin, Germany / Cologne [43] | Anti-S1 medium  (1.2–4.0), N (%) | 6-7 months | 6-7 months | Any persistent symptoms | 123 | 53 (44.5) | NA | NR |
| Augustin, Germany / Cologne [43] | Anti-S1 medium  (1.2–4.0), N (%) | 6-7 months | 6-7 months | No persistent symptoms | 230 | 76 (34.7) | NA | NR |
| Augustin, Germany / Cologne [43] | Anti-S1 high (>4.0),  N (%) | 6-7 months | 6-7 months | Any persistent symptoms | 123 | 45 (37.8) | NA | NR |
| Augustin, Germany / Cologne [43] | Anti-S1 high (>4.0),  N (%) | 6-7 months | 6-7 months | No persistent symptoms | 230 | 115 (52.5) | NA | NR |
| Augustin, Germany / Cologne [43] | OR, 95% CI, low (anti-S1 ≤ 1.1) compared to high (anti-S1 > 4.0) | 6-7 months | 6-7 months | Any persistent symptoms vs no persistent symptoms | 353 | 1.9  (0.99, 3.72) | NA | No increase / decrease |
| Augustin, Germany / Cologne [43] | OR, 95% CI, low (anti-S1 ≤ 1.1) compared to high (anti-S1 > 4.0) | 6-7 months | 6-7 months | Any persistent symptoms vs no persistent symptoms | 353 | 2.1  (0.99, 4.22) | Sex, age, preconditions, acute duration, acute symptom number, acute symptoms | No increase / decrease |
| Augustin, Germany / Cologne [43] | OR, 95% CI, medium (anti-S1 1.2 - 4) compared to high (anti-S1 > 4.0) | 6-7 months | 6-7 months | Any persistent symptoms vs no persistent symptoms | 353 | 1.8  (1.09, 2.91) | NA | Decrease |
| Augustin, Germany / Cologne [43] | OR, 95% CI, medium (anti-S1 1.2 - 4) compared to high (anti-S1 > 4.0) | 6-7 months | 6-7 months | Any persistent symptoms vs no persistent symptoms | 353 | 1.9  (1.13, 3.18) | Sex, age, preconditions, acute duration, acute symptom number, acute symptoms | Decrease |
| Lier, Saxony / Germany [63] | S/CO-Ratio, median (IQR) anti-RBD | 6-9 months | 6 months | Neuropsychiatric phenotype | 105 | 2,250 (9,897) | NA | Decrease |
| Lier, Saxony / Germany [63] | (S/CO-Ratio, median (IQR) anti-RBD | 6-9 months | 6 months | Without phenotype | 55 | 5,256 (12,518) | NA | Decrease |
| **Mixed (studies = 12; participants = 1216)** | | | | | | | | |
| Blomberg, Bergen / Norway [58] | RR (95% CI), anti-Spike | 2 months | 6 months | Fatigue score  (0-33) | 293 | 1.1  (1.07, 1.16) | NA | Increase |
| Blomberg, Bergen / Norway [58] | RR (95% CI), anti-Spike | 2 months | 6 months | Fatigue score  (0-33) | 293 | 1.1  (1.02, 1.12) | Sex, age, BMI, asthma/COPD, hypertension, chronic heart disease, rheumatic disease, diabetes, severity of acute illness, days in hospital | Increase |
| Blomberg, Bergen / Norway [58] | Mean (95% CI), anti-Spike | 2 months | 6 months | Fatigue | 108 | 4.1  (3.90, 4.20) | NA | Increase |
| Blomberg, Bergen / Norway [58] | Mean (95% CI), anti-Spike | 2 months | 6 months | No fatigue | 185 | 3.7  (3.60, 3.90) | NA | Increase |
| Blomberg, Bergen / Norway [58] | OR (95% CI), anti-Spike | 2 months | 6 months | Fatigue vs no fatigue | 293 | 1.7  (1.25, 2.39) | NA | Increase |
| Blomberg, Bergen / Norway [58] | OR (95% CI), anti-Spike | 2 months | 6 months | Fatigue vs no fatigue | 293 | 1.2  (0.81, 1.92) | Sex, age, BMI, asthma/COPD, chronic heart disease, severity of initial illness, days in hospital, antibiotic use, acute dyspnea, and acute fever | No increase / decrease |
| Durstenfeld, San Francisco / USA [66] | Median anti-RBD, IQR, (μg/ml ) | 3-4 months | 7-8 months | Dyspnea, chest pain, or palpitations | NR | 5.1 (8.5) | NA | Increase |
| Durstenfeld, San Francisco / USA [66] | Median anti-RBD, IQR, (μg/ml ) | 3-4 months | 7-8 months | No dyspnea, chest pain, or palpitations | NR | 2.9 (4.8) | NA | Increase |
| Durstenfeld, San Francisco / USA [66] | OR (95% CI) per doubling of anti-RBD antibody levels | 3-4 months | 7-8 months | Dyspnea, chest pain, or palpitations vs no dyspnea, chest pain, or palpitations | 73 | 1.4  (1.08, 1.81) | NA | Increase |
| Durstenfeld, San Francisco / USA [66] | OR (95% CI) per doubling of anti-RBD antibody levels | 3-4 months | 7-8 months | Dyspnea, chest pain, or palpitations vs no dyspnea, chest pain, or palpitations | 73 | 1.4  (1.06, 1.90) | Age, sex, hospitalization, time since symptom onset, HIV, and autoimmune disease | Increase |
| Durstenfeld, San Francisco / USA [66] | OR (95% CI) per doubling of anti-RBD antibody levels | 3-4 months | 7-8 months | Two or more cardiopulmonary symptoms vs other participants | 73 | 1.4  (0.96, 2.01) | NA | No increase / decrease |
| Gerhards, Mannheim / Germany [44] | Mean anti-RBD/S1 (SD), U/ml | Up to 8 months | 6 months | Any persistent symptoms | 22 | 299.3 (548.7) | NA | No increase / decrease |
| Gerhards, Mannheim / Germany [44] | Mean anti-RBD/S1 (SD), U/ml | Up to 8 months | 6 months | No persistent symptoms | 25 | 354.6 (408.9) | NA | No increase / decrease |
| Horton, New Jersey / USA [11] | Average anti-RBD levels over time vs days post COVID-19 positive | 5-6 months | ≥ 4 months post infection | All COVID-19 infected participants | 93 | Not specified | NA | Increase |
| Jia, Stanford / California, USA [45] | Hazard Ratio  (95% CI), anti-RBD | In the first month of illness | 6 months | Percentage of participants with resolved symptoms vs days post COVID-19 diagnosis | 105 | 2.1  (0.7, 6.5) | NA | No increase / decrease |
| Jia, Stanford / California, USA [45] | Hazard Ratio  (95% CI), anti-Spike | In the first month of illness | 6 months | Percentage of participants with resolved symptoms vs days post COVID-19 diagnosis | 105 | 2.1  (0.7, 6.7) | NA | No increase / decrease |
| Molnar, Hungary / Pecs [65] | Median anti-Spike, IQR, (U/mL) | > 3 months | > 3 months | Severe fatigue | 63 | 38.0 (97.0) | NA | Decrease |
| Molnar, Hungary / Pecs [65] | Median anti-Spike, IQR, (U/mL) | > 3 months | > 3 months | Less severe fatigue | 38 | 114.0 (653.0) | NA | Decrease |
| Peluso, San Francisco / California [50] | Mean anti-RBD (ug/mL), IQR | 3-4 months | 3-4 months | Any persistent symptoms | 73 | 4.04 (7.28) | NA | No increase / decrease |
| Peluso, San Francisco / California [50] | Mean anti-RBD (ug/mL), IQR | 3-4 months | 3-4 months | No persistent symptoms | 48 | 2.61 (5.52) | NA | No increase / decrease |
| Peluso, San Francisco / California [50] | Fold change in mean anti-RBD, 95% CI | 3-4 months | 3-4 months | Any persistent symptoms vs no persistent symptoms | 121 | 1.29  (0.73, 2.26) | NA | No increase / decrease |
| Peluso, San Francisco / California [50] | Fold change in mean anti-RBD, 95% CI | 3-4 months | 3-4 months | Any persistent symptoms vs no persistent symptoms | 121 | 1.10  (0.66, 1.81) | Adjusted for age, sex, and hospitalization status | No increase / decrease |
| Peluso, San Francisco / California [50] | Fold change in mean anti-RBD, 95% CI | 3-4 months | 3-4 months | Any persistent symptoms vs no persistent symptoms | 121 | 1.16  (0.65, 2.07) | Adjusted for age, sex, hospitalization status, history of autoimmune disease, and body mass index | No increase / decrease |
| Peluso, San Francisco / California [50] | Fold change in mean anti-RBD, 95% CI | 3-4 months | 3-4 months | Severe PCC – Top 25% of symptom number reported at late timepoint | 121 | 1.33  (0.76, 2.35) | NA | No increase / decrease |
| Peluso, San Francisco / California [64] | Geometric mean anti-RBD with interval [exp(log(geometric mean +/- one standard error))], ug/mL | 1-2 months | 3-4 months | With CNS PCC | 52 | 3.7  (3.4, 4.1) | NA | No increase / decrease |
| Peluso, San Francisco / California [64] | Geometric mean anti-RBD with interval [exp(log(geometric mean +/- one standard error))], ug/mL | 1-2 months | 3-4 months | Without CNS PCC | 69 | 3.1  (2.9, 3.4) | NA | No increase / decrease |
| Peluso, San Francisco / California [64] | Geometric mean anti-RBD with interval [exp(log(geometric mean +/- one standard error))], ug/mL | 3-4 months | 3-4 months | With CNS PCC | 52 | 3.0  (2.7, 3.3) | NA | No increase / decrease |
| Peluso, San Francisco / California [64] | Geometric mean anti-RBD with interval [exp(log(geometric mean +/- one standard error))], ug/mL | 3-4 months | 3-4 months | Without CNS PCC | 69 | 3.2  (2.9, 3.4) | NA | No increase / decrease |
| Peluso, San Francisco / California [64] | Fold change in mean anti-RBD, 95% CI | 1-2 months | 3-4 months | With CNS PCC vs no CNS PCC | 121 | 1.07  (0.60, 1.90) | NA | No increase / decrease |
| Peluso, San Francisco / California [64] | Fold change in mean anti-RBD, 95% CI | 3-4 months | 3-4 months | With CNS PCC vs no CNS PCC | 121 | 1.11  (0.63, 1.93) | NA | No increase / decrease |
| Peluso, San Francisco / California [64] | Fold change in mean anti-RBD, 95% CI | 1-2 months | 3-4 months | With CNS PCC vs no CNS PCC | 121 | 1.04  (0.59, 1.84) | Age | No increase / decrease |
| Peluso, San Francisco / California [64] | Fold change in mean anti-RBD, 95% CI | 3-4 months | 3-4 months | With CNS PCC vs no CNS PCC | 121 | 1.08  (0.62, 1.88) | Age | No increase / decrease |
| Peluso, San Francisco / California [64] | Fold change in mean anti-RBD, 95% CI | 1-2 months | 3-4 months | With CNS PCC vs no CNS PCC | 121 | 0.95  (0.56, 1.60) | Age, sex, and hospitalization | No increase / decrease |
| Peluso, San Francisco / California [64] | Fold change in mean anti-RBD, 95% CI | 3-4 months | 3-4 months | With CNS PCC vs no CNS PCC | 121 | 0.98  (0.59, 1.62) | Age, sex, and hospitalization | No increase / decrease |
| Peluso, San Francisco / California [64] | Fold change in mean anti-RBD, 95% CI | 1-2 months | 3-4 months | With CNS PCC vs no CNS PCC | 121 | 1.01  (0.56, 1.85) | Age, sex, hospitalization, history of autoimmune disease | No increase / decrease |
| Peluso, San Francisco / California [64] | Fold change in mean anti-RBD, 95% CI | 3-4 months | 3-4 months | With CNS PCC vs no CNS PCC | 121 | 1.05  (0.58, 1.92) | Age, sex, hospitalization, history of autoimmune disease | No increase / decrease |
| Peluso, San Francisco / California [64] | Fold change in mean anti-RBD, 95% CI | 1-2 months | 3-4 months | With CNS PCC vs no CNS PCC | 121 | 0.83  (0.45, 1.54) | Age, sex, hospitalization, history of autoimmune disease, BMI | No increase / decrease |
| Peluso, San Francisco / California [64] | Fold change in mean anti-RBD, 95% CI | 3-4 months | 3-4 months | With CNS PCC vs no CNS PCC | 121 | 0.87  (0.47, 1.61) | Age, sex, hospitalization, history of autoimmune disease, BMI | No increase / decrease |
| Peluso, San Francisco / California [64] | Fold change in mean anti-RBD, 95% CI | 1-2 months | 3-4 months | Any neuro PCC vs without neuro PCC | 121 | 1.06  (0.60, 1.87) | NA | No increase / decrease |
| Peluso, San Francisco / California [64] | Fold change in mean anti-RBD, 95% CI | 3-4 months | 3-4 months | Any neuro PCC vs without neuro PCC | 121 | 1.22  (0.70, 2.11) | NA | No increase / decrease |
| Peluso, San Francisco / California [64] | Fold change in mean anti-RBD, 95% CI | 1-2 months | 3-4 months | Any CNS symptom vs no PCC | 121 | 1.14  (0.61, 2.11) | NA | No increase / decrease |
| Peluso, San Francisco / California [64] | Fold change in mean anti-RBD, 95% CI | 3-4 months | 3-4 months | Any CNS symptom vs no PCC | 121 | 1.23  (0.67, 2.27) | NA | No increase / decrease |
| Peluso, San Francisco / California [49] | Differences in median anti-Spike (RU) at T1 and T4 | 1-2 months to 4-5 months | 4 months | Any persistent symptoms vs no persistent symptoms | 66 | Not specified | NA | No increase / decrease |
| Peluso, San Francisco / California [49] | Differences in median anti-RBD (RU) at T1 and T4 | 1-2 months to 4-5 months | 4 months | Any persistent symptoms vs no persistent symptoms | 66 | Not specified | NA | No increase / decrease |
| Sonnweber, Tyrol / Austria [54] | Anti-S1/S2 IgG, OR (95% CI), BAU, Quartile 1 | 2 months | 6 months | Any persistent symptoms vs no persistent symptoms | 134 | 1.0  (0.48, 2.28) | NA | No increase / decrease |
| Sonnweber, Tyrol / Austria [54] | Anti-S1/S2 IgG, OR (95% CI), BAU, Quartile 2 | 2 months | 6 months | Any persistent symptoms vs no persistent symptoms | 134 | 1.1  (0.51, 2.50) | NA | No increase / decrease |
| Sonnweber, Tyrol / Austria [54] | Anti-S1/S2 IgG, OR (95% CI), BAU, Quartile 3 | 2 months | 6 months | Any persistent symptoms vs no persistent symptoms | 134 | 0.7  (0.29, 1.41) | NA | No increase / decrease |
| Sonnweber, Tyrol / Austria [54] | Anti-S1/S2 IgG, OR (95% CI), BAU, Quartile 4 | 2 months | 6 months | Any persistent symptoms vs no persistent symptoms | 134 | 1.3  (0.60, 2.95) | NA | No increase / decrease |
| Varnai, Hungary / Pecs [30] | Median anti-Spike, IQR, (U/mL) | 6-7 months | 6-7 months | Severe fatigue | 57 | 3,723 (10,021) | NA | No increase / decrease |
| Varnai, Hungary / Pecs [30] | Median anti-Spike, IQR, (U/mL) | 6-7 months | 6-7 months | Non-severe fatigue | 50 | 6,949 (11,070) | NA | No increase / decrease |
| Wynberg, Amsterdam, The Netherlands [55] | Anti-Spike, median difference in posterior means (95% CrI) | 1-2 months | 3 months | Any persistent symptoms vs no persistent symptoms | 316 | 0.11  (-0.10, 0.34) | NA | No increase / decrease |
| Wynberg, Amsterdam, The Netherlands [55] | Anti-RBD, median difference in posterior means (95% CrI) | 1-2 months | 3 months | Any persistent symptoms vs no persistent symptoms | 316 | 0.09  (-0.08, 0.32) | NA | No increase / decrease |
| Wynberg, Amsterdam, The Netherlands [55] | Median half-life of spike-binding IgG  (95% CrI) | 1-2 months post onset | 3 months | Any persistent symptoms | 186 | 233  (183, 324) | NA | No increase / decrease |
| Wynberg, Amsterdam, The Netherlands [55] | Median half-life of spike-binding IgG  (95% CrI) | 1-2 months post onset | 3 months | No persistent symptoms | 130 | 170  (125, 252) | NA | No increase / decrease |
| Wynberg, Amsterdam, The Netherlands [55] | Median half-life of RBD-binding IgG  (95% CrI) | 1-2 months post onset | 3 months | Any persistent symptoms | 186 | 181  (147, 230) | NA | No increase / decrease |
| Wynberg, Amsterdam, The Netherlands [55] | Median half-life of RBD-binding IgG  (95% CrI) | 1-2 months post onset | 3 months | No persistent symptoms | 130 | 144  (113, 196) | NA | No increase / decrease |
| ^a^Trend as reported by study authors  CSQ - COVID-19 symptoms questionnaire; IQR - interquartile range; RU - relative light units; S/CO - signal to cut-off ratio; NA - Not applicable; NR - Not reported; T1 - Visit 1; T4 - Visit 4; OR - odds ratio; CI - confidence interval; RBD - receptor binding domain; BAU - binding antigen unit; CrI - credible interval | | | | | | | | |

| **Table S3: Neutralizing antibody comparisons among groups with and without persistent symptoms ≥12 weeks post COVID-19, stratified by level of care requirements during the acute phase of illness** | | | | | | | | |
| --- | --- | --- | --- | --- | --- | --- | --- | --- |
| **Study cohort** | **Measure / unit** | **Timepoints assessed** | | **Comparisons** | | | | **Overall trend^a^** |
|  |  | **Serology** | **Post-acute** | **Group** | **Participants assessed (N)** | **Results** | **Variables adjusted** |  |
| **Hospitalized (studies = 2; participants = 193)** | | | | | | | | |
| García-Abellán, Alicante / Spain [28] | Median (IQR) NeutraLISA,% IH | 12 months | 6 and 12 months | Highest CSQ scores after discharge | 14 | 27.3 (59.0) | NA | No increase / decrease |
| García-Abellán, Alicante / Spain [28] | Median (IQR) NeutraLISA,% IH | 12 months | 6 and 12 months | Other participants | 58 | 69.7 (44.0) | NA | No increase / decrease |
| García-Abellán, Alicante / Spain [28] | NeutraLISA positive,  N (%) | 12 months | 6 and 12 months | Highest CSQ scores after discharge | 14 | 6.0 (42.9) | NA | Decrease |
| García-Abellán, Alicante / Spain [28] | NeutraLISA positive,  N (%) | 12 months | 6 and 12 months | Other participants | 58 | 46.0 (79.3) | NA | Decrease |
| García-Abellán, Alicante / Spain [28] | aHR (95% CI) NeutraLISA positive, AU/Ml | 12 months | 6 and 12 months | Highest CSQ scores after discharge vs other participants | 72 | 0.98  (0.97, 0.99) | Sex and ICU stay | Decrease |
| Zhan, Xiangyang / China [56] | Median 50% pseudovirus neutralization titers (pNT50) | 10-12 months | 10-12 months | Any persistent symptoms | 36 | 18.8 | NA | Decrease |
| Zhan, Xiangyang / China [56] | Median 50% pseudovirus neutralization titers (pNT50) | 10-12 months | 10-12 months | No persistent symptoms | 85 | 29.2 | NA | Decrease |
| **Non-hospitalized (studies = 1; participants = 160)** | | | | | | | | |
| Lier, Saxony / Germany [63] | S/CO, median (IQR), neutralizing antibodies | 6-9 months | 6 months | Neuropsychiatric phenotype | 105 | 465 (1,398) | NA | Decrease |
| Lier, Saxony / Germany [63] | S/CO, median (IQR), neutralizing antibodies | 6-9 months | 6 months | Without phenotype | 55 | 746 (1,539) | NA | Decrease |
| **Mixed (studies = 4; participants = 771)** | | | | | | | | |
| Blomberg, Bergen / Norway [58] | Microneutralizing titres RR (95% CI) | 2 months | 6 months | Fatigue score  (0-33) | 293 | 1.1  (1.08-1.19) | Sex, age, BMI, asthma/COPD, hypertension, chronic heart disease, rheumatic disease, diabetes, severity of acute illness, days in hospital | Increase |
| Blomberg, Bergen / Norway [58] | Mean microneutralizing titres (95% CI) | 2 months | 6 months | Fatigue | 108 | 2.2  (2.10, 2.40) | NA | Increase |
| Blomberg, Bergen / Norway [58] | Mean microneutralizing titres (95% CI) | 2 months | 6 months | No fatigue | 185 | 1.9  (1.80, 2.00) | NA | Increase |
| Blomberg, Bergen / Norway [58] | Microneutralizing titres RR (95% CI) | 2 months | 6 months | Fatigue vs no fatigue | 293 | 1.8  (1.27, 2.52) | NA | Increase |
| Peluso, San Francisco / California [49] | Differences in median neutralizing capacity (infectious dose, 50% [ID50]) at T1 and T4 | 1-2 months to 4-5 months | 4 months | Any persistent symptoms vs no persistent symptoms | 66 | Not specified | NA | No increase / decrease |
| Seeßle, Heidelberg / Germany [52] | Median relative S1-ACE2 competition efficiency (%): Group differences based on Mann-Whitney U test | 5, 9, and 12 months | 12 months | Any persistent symptoms vs no persistent symptoms | 96 | Not specified | NA | No increase / decrease |
| Wynberg, Amsterdam, The Netherlands [55] | Neutralizing antibodies, median difference in posterior means (95% credible interval) | 1-2 months | 3 months | Any persistent symptoms vs no persistent symptoms | 316 | 0.17  (-0.05, 0.40) | NA | No increase / decrease |
| ^a^Trend as reported by study authors  CSQ - COVID-19 symptoms questionnaire; IQR - interquartile range; RU - relative light units; S/CO - signal to cut-off ratio; NA - Not applicable; NR - Not reported; T1 - Visit 1; T4 - Visit 4; OR - odds ratio; CI - confidence interval; IH – inhibition; aHR – adjusted hazard ratio; RR - risk ratio | | | | | | | | |

| **Table S4: Anti-N IgG comparisons among groups with and without persistent symptoms ≥12 weeks post COVID-19, stratified by time interval (months) between COVID-19 infection and serological sampling** | | | | | | | |
| --- | --- | --- | --- | --- | --- | --- | --- |
| **Study cohort** | **LOC** | **Measure / unit** | **Comparisons** | | | | **Overall trend^a^** |
|  |  |  | **Group** | **Participants assessed (N)** | **Results** | **Variables adjusted** |  |
| **Serological sampling < 3 months post COVID-19 (studies = 5; participants = 412)** | | | | | | | |
| Bilich, Tübingen / Germany [42] | NR | Median change in  anti-N titres (IQR) | Any persistent symptoms | 14 | 6.5 (38.6) | NA | No increase / decrease |
| Bilich, Tübingen / Germany [42] | NR | Median change in  anti-N titres (IQR) | No persistent symptoms | 37 | 1.2 (12.6) | NA | No increase / decrease |
| García-Abellán, Alicante / Spain [60] | Hospitalized | N (%) seropositive | Highest CSQ scores after discharge | 28 | 17 (68.0) | NA | No increase / decrease |
| García-Abellán, Alicante / Spain [60] | Hospitalized | N (%) seropositive | Other participants | 88 | 59 (74.0) | NA | No increase / decrease |
| García-Abellán, Alicante / Spain [60] | Hospitalized | N (%) seropositive | Highest CSQ scores after discharge | 28 | 17 (68.0) | NA | No increase / decrease |
| García-Abellán, Alicante / Spain [60] | Hospitalized | N (%) seropositive | Other participants | 88 | 61 (72.6) | NA | No increase / decrease |
| Jia, Stanford / California, USA [45] | Mixed | N (%), (High > 149,452 AU/mL) | Probability of reporting persistent symptoms | NR | 36.0 % | NA | No increase / decrease |
| Jia, Stanford / California, USA [45] | Mixed | N (%), (Low <=149,452 AU/ml) | Probability of reporting persistent symptoms | NR | 55.0 % | NA | No increase / decrease |
| Jia, Stanford / California, USA [45] | Mixed | Hazard Ratio  (95% CI) | Percentage of participants with resolved symptoms vs days post COVID-19 diagnosis | 105 | 3.5 (1.0, 11.6) | NA | Decrease |
| Pilmis, Paris / France [51] | Non-hospitalized | Inter-group comparisons using the Mann-Whitney test | Any participants vs no participants | 74 | Not specified | NA | No increase / decrease |
| Peluso, San Francisco / California [49] | Mixed | Differences in median anti-N (RU) at T1 and T4 | Any persistent symptoms vs no persistent symptoms | 66 | Not specified | NA | No increase / decrease |
| **Serological sampling 3 – <6 months post COVID-19 (studies = 4; participants = 407)** | | | | | | | |
| Bilich, Tübingen / Germany [42] | NR | Median anti-N titres (IQR) | Any persistent symptoms | 14 | 58.1 (50.6) | NA | Increase |
| Bilich, Tübingen / Germany [42] | NR | Median anti-N titres (IQR) | No persistent symptoms | 37 | 19.7 (55.7) | NA | Increase |
| Bilich, Tübingen / Germany [42] | NR | Median change in  anti-N titres (IQR) | Any persistent symptoms | 14 | 6.5 (38.6) | NA | No increase / decrease |
| Bilich, Tübingen / Germany [42] | NR | Median change in  anti-N titres (IQR) | No persistent symptoms | 37 | 1.2 (12.6) | NA | No increase / decrease |
| Molnar, Hungary / Pecs [65] | Mixed | Median (IQR), (U/mL) | Severe fatigue | 63 | 24.8 (50.0) | NA | Decrease |
| Molnar, Hungary / Pecs [65] | Mixed | Median (IQR), (U/mL) | Less severe fatigue | 38 | 90.0 (60.0) | NA | Decrease |
| Sneller, Maryland / USA [53] | Mixed | N (%) | Any persistent symptoms | 104 | 90 (86.5) | NA | No increase / decrease |
| Sneller, Maryland / USA [53] | Mixed | N (%) | No persistent symptoms | 85 | 69 (81.2) | NA | No increase / decrease |
| Peluso, San Francisco / California [49] | Mixed | Differences in median anti-N (RU) at T1 and T4 | Any persistent symptoms vs no persistent symptoms | 66 | NR | NA | No increase / decrease |
| **Serological sampling 6 – <9 months post COVID-19 (studies = 4; participants = 457)** | | | | | | | |
| García-Abellán, Alicante / Spain [60] | Hospitalized | N (%) seropositive | Highest CSQ scores after discharge | 28 | 19 (73.1) | NA | No increase / decrease |
| García-Abellán, Alicante / Spain [60] | Hospitalized | N (%) seropositive | Other participants | 88 | 61 (71.8) | NA | No increase / decrease |
| García-Abellán, Alicante / Spain [60] | Hospitalized | Peak serum concentration (S/CO) | Highest CSQ scores after discharge | 28 | 3.8 (2.9) | NA | No increase / decrease |
| García-Abellán, Alicante / Spain [60] | Hospitalized | Peak serum concentration (S/CO) | Other participants | 88 | 4.1 (2.2) | NA | No increase / decrease |
| Lier, Saxony / Germany [63] | Non-hospitalized | Median (IQR), (S/CO) | Neuropsychiatric phenotype | 105 | 1.4 (1.2) | NA | No increase / decrease |
| Lier, Saxony / Germany [63] | Non-hospitalized | Median (IQR), (S/CO) | Without phenotype | 55 | 1.4 (1.3) | NA | No increase / decrease |
| Pilmis, Paris / France [51] | Non-hospitalized | Inter-group comparisons using the Mann-Whitney test | Any persistent symptoms vs no persistent symptoms | 74 | Not specified | NA | No increase / decrease |
| Varnai, Hungary / Pecs [30] | Mixed | Median (IQR), (U/mL) | Severe fatigue | 57 | 27.0 (75.0) | NA | Decrease |
| Varnai, Hungary / Pecs [30] | Mixed | Median (IQR), (U/mL) | Non-severe fatigue | 50 | 98.0 (123.0) | NA | Decrease |
| **Serological sampling 9 – 12 months post COVID-19 (studies = 1; participants = 121)** | | | | | | | |
| Zhan, Xiangyang / China [56] | Hospitalized | Beta (95% CI) | Any persistent symptoms vs no persistent symptoms | 121 | -0.16 (-0.34, 0.01) | Time from discharge, length of stay, age, sex, severe disease, glucocorticoids, interferons | No increase / decrease |
| ^a^Trend as reported by study authors  CSQ - COVID-19 symptoms questionnaire; IQR - interquartile range; RU - relative light units; S/CO - signal to cut-off ratio; NA - Not applicable; NR - Not reported; T1 - Visit 1; T4 - Visit 4; OR - odds ratio; CI - confidence interval | | | | | | | |

| **Table S5: Anti-Spike IgG comparisons (Spike protein, subunits, or RBD) among groups with and without persistent symptoms ≥12 weeks post COVID-19, stratified by time interval (months) between COVID-19 infection and serological sampling** | | | | | | | | |
| --- | --- | --- | --- | --- | --- | --- | --- | --- |
| **Study cohort** | **LOC** | **Measure / unit** |  | **Comparisons** | | | | **Overall trend^a^** |
|  |  |  | **Group** | | **Participants assessed (N)** | **Results** | **Variables adjusted** |  |
| **Serological sampling < 3 months post COVID-19 (studies = 10; participants = 2038)** | | | | | | | | |
| Augustin, Germany / Cologne [43] | Non-hospitalized | S/CO-Ratio, median (IQR) anti-S1 | Any persistent symptoms | | 123 | 3 (5.0) | NA | NR |
| Augustin, Germany / Cologne [43] | Non-hospitalized | S/CO-Ratio, median (IQR) anti-S1 | No persistent symptoms | | 230 | 4 (5.0) | NA | NR |
| Bilich, Tübingen / Germany [42] | NR | Median change in anti-S1 titres (IQR) | Any persistent symptoms | | 14 | -1.9 (-2.3) | NA | No increase / decrease |
| Bilich, Tübingen / Germany [42] | NR | Median change in anti-S1 titres (IQR) | No persistent symptoms | | 37 | -0.6 (-2.2) | NA | No increase / decrease |
| Blomberg, Bergen / Norway [58] | Mixed | RR (95% CI), anti-Spike | Fatigue score  (0-33) | | 293 | 1.1  (1.07, 1.16) | NA | Increase |
| Blomberg, Bergen / Norway [58] | Mixed | RR (95% CI), anti-Spike | Fatigue score  (0-33) | | 293 | 1.1  (1.02, 1.12) | Sex, age, BMI, asthma/COPD, hypertension, chronic heart disease, rheumatic disease, diabetes, severity of acute illness, days in hospital | Increase |
| Blomberg, Bergen / Norway [58] | Mixed | Mean  (95% CI), anti-Spike | Fatigue | | 108 | 4.1  (3.90, 4.20) | NA | Increase |
| Blomberg, Bergen / Norway [58] | Mixed | Mean  (95% CI), anti-Spike | No fatigue | | 185 | 3.7  (3.60, 3.90) | NA | Increase |
| Blomberg, Bergen / Norway [58] | Mixed | OR (95% CI), anti-Spike | Fatigue vs no fatigue | | 293 | 1.7  (1.25, 2.39) | NA | Increase |
| Blomberg, Bergen / Norway [58] | Mixed | OR (95% CI), anti-Spike | Fatigue vs no fatigue | | 293 | 1.2  (0.81, 1.92) | Sex, age, BMI, asthma/COPD, chronic heart disease, severity of initial illness, days in hospital, antibiotic use, acute dyspnea, and acute fever | No increase / decrease |
| García-Abellán, Alicante / Spain [60] | Hospitalized | N (%) IgG-Spike seropositive | Highest CSQ scores after discharge | | 28 | 17 (68.0) | NA | No increase / decrease |
| García-Abellán, Alicante / Spain [60] | Hospitalized | N (%) IgG-Spike seropositive | Other participants | | 88 | 58 (72.5) | NA | No increase / decrease |
| García-Abellán, Alicante / Spain [60] | Hospitalized | N (%) IgG-Spike seropositive | Highest CSQ scores after discharge | | 28 | 17 (68.0) | NA | No increase / decrease |
| García-Abellán, Alicante / Spain [60] | Hospitalized | N (%) IgG-Spike seropositive | Other participants | | 88 | 61 (72.6) | NA | No increase / decrease |
| Jia, Stanford / California, USA [45] | Mixed | Hazard Ratio  (95% CI), anti-RBD | Percentage of participants with resolved symptoms vs days post COVID-19 diagnosis | | 105 | 2.1  (0.7, 6.5) | NA | No increase / decrease |
| Jia, Stanford / California, USA [45] | Mixed | Hazard Ratio  (95% CI), anti-Spike | Percentage of participants with resolved symptoms vs days post COVID-19 diagnosis | | 105 | 2.1  (0.7, 6.7) | NA | No increase / decrease |
| Ozonoff, Boston / USA [46] | Hospitalized | Anti-RBD over time: trend lines representing the fit of a generalized additive model (GAM) | Any persistent symptoms vs no persistent symptoms | | 589 | No specified | NA | No increase / decrease |
| Peluso, San Francisco / California [64] | Mixed | Geometric mean anti-RBD with interval [exp(log(geometric mean +/- one standard error))], ug/mL | With CNS PCC | | 52 | 3.7 (3.4, 4.1) | NA | No increase / decrease |
| Peluso, San Francisco / California [64] | Mixed | Geometric mean anti-RBD with interval [exp(log(geometric mean +/- one standard error))], ug/mL | Without CNS PCC | | 69 | 3.1 (2.9, 3.4) | NA | No increase / decrease |
| Peluso, San Francisco / California [64] | Mixed | Fold change in mean anti-RBD, 95% CI | With CNS PCC vs no CNS PCC | | 121 | 1.07  (0.60, 1.90) | NA | No increase / decrease |
| Peluso, San Francisco / California [64] | Mixed | Fold change in mean anti-RBD, 95% CI | With CNS PCC vs no CNS PCC | | 121 | 1.04  (0.59, 1.84) | Age | No increase / decrease |
| Peluso, San Francisco / California [64] | Mixed | Fold change in mean anti-RBD, 95% CI | With CNS PCC vs no CNS PCC | | 121 | 0.95  (0.56, 1.60) | Age, sex, and hospitalization | No increase / decrease |
| Peluso, San Francisco / California [64] | Mixed | Fold change in mean anti-RBD, 95% CI | With CNS PCC vs no CNS PCC | | 121 | 1.01  (0.56, 1.85) | Age, sex, hospitalization, history of autoimmune disease | No increase / decrease |
| Peluso, San Francisco / California [64] | Mixed | Fold change in mean anti-RBD, 95% CI | With CNS PCC vs no CNS PCC | | 121 | 0.83  (0.45, 1.54) | Age, sex, hospitalization, history of autoimmune disease, BMI | No increase / decrease |
| Peluso, San Francisco / California [64] | Mixed | Fold change in mean anti-RBD, 95% CI | Any neuro PCC vs without neuro PCC | | 121 | 1.06  (0.60, 1.87) | NA | No increase / decrease |
| Peluso, San Francisco / California [64] | Mixed | Fold change in mean anti-RBD, 95% CI | Any CNS symptom vs no PCC | | 121 | 1.14  (0.61, 2.11) | NA | No increase / decrease |
| Peluso, San Francisco / California [49] | Mixed | Differences in median anti-Spike (RU) at T1 and T4 | Any persistent symptoms vs no persistent symptoms | | 66 | NR | NA | No increase / decrease |
| Peluso, San Francisco / California [49] | Mixed | Differences in median anti-RBD (RU) at T1 and T4 | Any persistent symptoms vs no persistent symptoms | | 66 | NR | NA | No increase / decrease |
| Sonnweber, Tyrol / Austria [54] | Mixed | Anti-S1/S2 IgG, OR (95% CI), BAU, Quartile 1 | Any persistent symptoms vs no persistent symptoms | | 134 | 1.0  (0.48, 2.28) | NA | No increase / decrease |
| Sonnweber, Tyrol / Austria [54] | Mixed | Anti-S1/S2 IgG, OR (95% CI), BAU, Quartile 2 | Any persistent symptoms vs no persistent symptoms | | 134 | 1.1  (0.51, 2.50) | NA | No increase / decrease |
| Sonnweber, Tyrol / Austria [54] | Mixed | Anti-S1/S2 IgG, OR (95% CI), BAU, Quartile 3 | Any persistent symptoms vs no persistent symptoms | | 134 | 0.7  (0.29, 1.41) | NA | No increase / decrease |
| Sonnweber, Tyrol / Austria [54] | Mixed | Anti-S1/S2 IgG, OR (95% CI), BAU, Quartile 4 | Any persistent symptoms vs no persistent symptoms | | 134 | 1.3  (0.60, 2.95) | NA | No increase / decrease |
| Wynberg, Amsterdam, The Netherlands [55] | Mixed | Anti-Spike, median difference in posterior means (95% credible interval) | Any persistent symptoms vs no persistent symptoms | | 316 | 0.11  (-0.10, 0.34) | NA | No increase / decrease |
| Wynberg, Amsterdam, The Netherlands [55] | Mixed | Anti-RBD, median difference in posterior means (95% credible interval) | Any persistent symptoms vs no persistent symptoms | | 316 | 0.09  (-0.08, 0.32) | NA | No increase / decrease |
| Wynberg, Amsterdam, The Netherlands [55] | Mixed | Median half-life of spike-binding IgG  (95% CrI) | Any persistent symptoms | | 186 | 233  (183, 324) | NA | No increase / decrease |
| Wynberg, Amsterdam, The Netherlands [55] | Mixed | Median half-life of spike-binding IgG  (95% CrI) | No persistent symptoms | | 130 | 170  (125, 252) | NA | No increase / decrease |
| Wynberg, Amsterdam, The Netherlands [55] | Mixed | Median half-life of RBD-binding IgG  (95% CrI) | Any persistent symptoms | | 186 | 181  (147, 230) | NA | No increase / decrease |
| Wynberg, Amsterdam, The Netherlands [55] | Mixed | Median half-life of RBD-binding IgG  (95% CrI) | No persistent symptoms | | 130 | 144  (113, 196) | NA | No increase / decrease |
| **Serological sampling 3 - < 6 months post COVID-19 (studies = 8; participants = 482)** | | | | | | | | |
| Bilich, Tübingen / Germany [42] | NR | Median anti-S1 titres (IQR) | Any persistent symptoms | | 14 | 2.5 (2.5) | NA | No increase / decrease |
| Bilich, Tübingen / Germany [42] | NR | Median anti-S1 titres (IQR) | No persistent symptoms | | 37 | 2.6 (3.0) | NA | No increase / decrease |
| Bilich, Tübingen / Germany [42] | NR | Median change in anti-S1 titres (IQR) | Any persistent symptoms | | 14 | -1.9 (-2.3) | NA | No increase / decrease |
| Bilich, Tübingen / Germany [42] | NR | Median change in anti-S1 titres (IQR) | No persistent symptoms | | 37 | -0.6 (-2.2) | NA | No increase / decrease |
| Horton,  New Jersey / USA [59] | Mixed | Average anti-RBD levels over time vs days post COVID-19 positive | All COVID-19 infected participants | | 93 | NR | NA | Increase |
| Molnar, Hungary / Pecs [65] | Mixed | Median anti-Spike, IQR, (U/mL) | Severe fatigue | | 63 | 38.0 (97.0) | NA | Decrease |
| Molnar, Hungary / Pecs [65] | Mixed | Median anti-Spike, IQR, (U/mL) | Less severe fatigue | | 38 | 114.0 (653.0) | NA | Decrease |
| Peluso, San Francisco / California [50] | Mixed | Mean anti-RBD (ug/mL), IQR | Any persistent symptoms | | 73 | 4.04 (7.28) | NA | No increase / decrease |
| Peluso, San Francisco / California [50] | Mixed | Mean anti-RBD (ug/mL), IQR | No persistent symptoms | | 48 | 2.61 (5.52) | NA | No increase / decrease |
| Peluso, San Francisco / California [50] | Mixed | Fold change in mean anti-RBD, 95% CI | Any persistent symptoms vs no persistent symptoms | | 121 | 1.3  (0.7, 2.3) | NA | No increase / decrease |
| Peluso, San Francisco / California [50] | Mixed | Fold change in mean anti-RBD, 95% CI | Any persistent symptoms vs no persistent symptoms | | 121 | 1.1  (0.7, 1.8) | Adjusted for age, sex, and hospitalization status | No increase / decrease |
| Peluso, San Francisco / California [50] | Mixed | Fold change in mean anti-RBD, 95% CI | Any persistent symptoms vs no persistent symptoms | | 121 | 1.2  (0.7, 2.1) | Adjusted for age, sex, hospitalization status, history of autoimmune disease, and body mass index | No increase / decrease |
| Peluso, San Francisco / California [50] | Mixed | Fold change in mean anti-RBD, 95% CI | Severe PCC – Top 25% of symptom number reported at late timepoint | | 121 | 1.3  (0.8, 2.4) | NA | No increase / decrease |
| Peluso, San Francisco / California [64] | Mixed | Geometric mean anti-RBD with interval [exp(log(geometric mean +/- one standard error))], ug/mL | With CNS PCC | | 52 | 3.0 (2.7, 3.3) | NA | No increase / decrease |
| Peluso, San Francisco / California [64] | Mixed | Geometric mean anti-RBD with interval [exp(log(geometric mean +/- one standard error))], ug/mL | Without CNS PCC | | 69 | 3.2 (2.9, 3.4) | NA | No increase / decrease |
| Peluso, San Francisco / California [64] | Mixed | Fold change in mean anti-RBD, 95% CI | With CNS PCC vs no CNS PCC | | 121 | 1.11  (0.63, 1.93) | NA | No increase / decrease |
| Peluso, San Francisco / California [64] | Mixed | Fold change in mean anti-RBD, 95% CI | With CNS PCC vs no CNS PCC | | 121 | 1.08  (0.62, 1.88) | Age | No increase / decrease |
| Peluso, San Francisco / California [64] | Mixed | Fold change in mean anti-RBD, 95% CI | With CNS PCC vs no CNS PCC | | 121 | 0.98  (0.59, 1.62) | Age, sex, and hospitalization | No increase / decrease |
| Peluso, San Francisco / California [64] | Mixed | Fold change in mean anti-RBD, 95% CI | With CNS PCC vs no CNS PCC | | 121 | 1.05  (0.58, 1.92) | Age, sex, hospitalization, history of autoimmune disease | No increase / decrease |
| Peluso, San Francisco / California [64] | Mixed | Fold change in mean anti-RBD, 95% CI | With CNS PCC vs no CNS PCC | | 121 | 0.87  (0.47, 1.61) | Age, sex, hospitalization, history of autoimmune disease, BMI | No increase / decrease |
| Peluso, San Francisco / California [64] | Mixed | Fold change in mean anti-RBD, 95% CI | Any neuro PCC vs without neuro PCC | | 121 | 1.22  (0.70, 2.11) | NA | No increase / decrease |
| Peluso, San Francisco / California [64] | Mixed | Fold change in mean anti-RBD, 95% CI | Any CNS symptom vs no PCC | | 121 | 1.23  (0.67, 2.27) | NA | No increase / decrease |
| Peluso, San Francisco / California [49] | Mixed | Differences in median anti-Spike (RU) at T1 and T4 | Any persistent symptoms vs no persistent symptoms | | 66 | NR | NA | No increase / decrease |
| Peluso, San Francisco / California [49] | Mixed | Differences in median anti-Spike (RU) at T1 and T4 | Any persistent symptoms vs no persistent symptoms | | 66 | NR | NA | No increase / decrease |
| García-Abellán, Alicante / Spain [60] | Hospitalized | Peak IgG-Spike, serum concentration - S/CO (IQR) | Highest CSQ scores after discharge vs other participants | | 116 | 0.9  (0.79, 0.99) | Age, sex, Charlson comorbidity index, WHO severity ordinal scale score, testing positive for SARS-CoV-2 RT-PCR at 1-month visit and tocilizumab use | Decrease |
| García-Abellán, Alicante / Spain [60] | Hospitalized | Peak IgG-Spike, serum concentration - S/CO (IQR) | Median CSQ score after discharge vs other participants | | 116 | 0.9  (0.78, 1.07) | Age, sex, Charlson comorbidity index, WHO severity ordinal scale score, testing positive for SARS-CoV-2 RT-PCR at 1-month visit and tocilizumab use | No increase / decrease |
| García-Abellán, Alicante / Spain [60] | Hospitalized | Peak IgG-Spike, serum concentration - S/CO (IQR) | Any symptoms after discharge vs other participants | | 116 | 1.0  (0.90, 1.20) | Age, sex, Charlson comorbidity index, WHO severity ordinal scale score, testing positive for SARS-CoV-2 RT-PCR at 1-month visit and tocilizumab use | No increase / decrease |
| Durstenfeld, San Francisco / USA [66] | Mixed | Median anti-RBD, IQR, (μg/ml) | Dyspnea, chest pain, or palpitations | | NR | 5.1 (8.5) | NA | Increase |
| Durstenfeld, San Francisco / USA [66] | Mixed | Median anti-RBD, IQR, (μg/ml) | No dyspnea, chest pain, or palpitations | | NR | 2.9 (4.8) | NA | Increase |
| Durstenfeld, San Francisco / USA [66] | Mixed | OR (95% CI) per doubling of anti-RBD antibody levels | Dyspnea, chest pain, or palpitations vs no dyspnea, chest pain, or palpitations | | 73 | 1.4  (1.08, 1.81) | NA | Increase |
| Durstenfeld, San Francisco / USA [66] | Mixed | OR (95% CI) per doubling of anti-RBD antibody levels | Dyspnea, chest pain, or palpitations vs no dyspnea, chest pain, or palpitations | | 73 | 1.4  (1.06, 1.90) | Age, sex, hospitalization, time since symptom onset, HIV, and autoimmune disease | Increase |
| Durstenfeld, San Francisco / USA [66] | OR (95% CI) per doubling of anti-RBD antibody levels | OR (95% CI) per doubling of anti-RBD antibody levels | Two or more cardiopulmonary symptoms vs other participants | | 73 | 1.4  (0.96, 2.01) | NA | No increase / decrease |
| **Serological sampling 6 - < 9 months post COVID-19 (studies = 5; participants = 783)** | | | | | | | | |
| Augustin, Germany / Cologne [43] | Non-hospitalized | S/CO-Ratio, median (IQR) anti-S1 | Any persistent symptoms | | 123 | 2 (3.0) | NA | NR |
| Augustin, Germany / Cologne [43] | Non-hospitalized | S/CO-Ratio, median (IQR) anti-S1 | No persistent symptoms | | 230 | 2 (3.0) | NA | NR |
| Augustin, Germany / Cologne [43] | Non-hospitalized | Anti-S1 low (≤1.1),  N (%) | Any persistent symptoms | | 123 | 21 (17.6) | NA | NR |
| Augustin, Germany / Cologne [43] | Non-hospitalized | Anti-S1 low (≤1.1),  N (%) | No persistent symptoms | | 230 | 28 (12.8) | NA | NR |
| Augustin, Germany / Cologne [43] | Non-hospitalized | Anti-S1 medium (1.2–4.0), N (%) | Any persistent symptoms | | 123 | 53 (44.5) | NA | NR |
| Augustin, Germany / Cologne [43] | Non-hospitalized | Anti-S1 medium (1.2–4.0), N (%) | No persistent symptoms | | 230 | 76 (34.7) | NA | NR |
| Augustin, Germany / Cologne [43] | Non-hospitalized | Anti-S1 high (>4.0),  N (%) | Any persistent symptoms | | 123 | 45 (37.8) | NA | NR |
| Augustin, Germany / Cologne [43] | Non-hospitalized | Anti-S1 high (>4.0),  N (%) | No persistent symptoms | | 230 | 115 (52.5) | NA | NR |
| Augustin, Germany / Cologne [43] | Non-hospitalized | OR, 95% CI, low (anti-S1 ≤ 1.1) compared to high (anti-S1 > 4.0) | Any persistent symptoms vs no persistent symptoms | | 353 | 1.9  (0.99, 3.72) | NA | No increase / decrease |
| Augustin, Germany / Cologne [43] | Non-hospitalized | OR, 95% CI, low (anti-S1 ≤ 1.1) compared to high (anti-S1 > 4.0) | Any persistent symptoms vs no persistent symptoms | | 353 | 2.1  (0.99, 4.22) | Sex, age, preconditions, acute duration, acute symptom number, acute symptoms | No increase / decrease |
| Augustin, Germany / Cologne [43] | Non-hospitalized | OR, 95% CI, medium (anti-S1 1.2 - 4) compared to high (anti-S1 > 4.0) | Any persistent symptoms vs no persistent symptoms | | 353 | 1.8  (1.09, 2.91) | NA | Decrease |
| Augustin, Germany / Cologne [43] | Non-hospitalized | OR, 95% CI, medium (anti-S1 1.2 - 4) compared to high (anti-S1 > 4.0) | Any persistent symptoms vs no persistent symptoms | | 353 | 1.9  (1.13, 3.18) | Sex, age, preconditions, acute duration, acute symptom number, acute symptoms | Decrease |
| García-Abellán, Alicante / Spain [60] | Hospitalized | N (%) IgG-Spike seropositive | Any persistent symptoms | | 28 | 19 (73.1) | NA | No increase / decrease |
| García-Abellán, Alicante / Spain [60] | Hospitalized | N (%) IgG-Spike seropositive | No persistent symptoms | | 88 | 62 (72.9) | NA | No increase / decrease |
| García-Abellán, Alicante / Spain [60] | Hospitalized | Peak IgG-Spike, serum concentration - S/CO (IQR) | Any persistent symptoms | | 28 | 5.1 (3.9) | NA | No increase / decrease |
| García-Abellán, Alicante / Spain [60] | Hospitalized | Peak IgG-Spike, serum concentration - S/CO (IQR) | No persistent symptoms | | 88 | 6.3 (4.6) | NA | No increase / decrease |
| García-Abellán, Alicante / Spain [60] | Hospitalized | Trough IgG-Spike, serum concentration - S/CO (IQR) | Any persistent symptoms | | 28 | 3.9 (28.0) | NA | No increase / decrease |
| García-Abellán, Alicante / Spain [60] | Hospitalized | Trough IgG-Spike, serum concentration - S/CO (IQR | No persistent symptoms | | 88 | 3.7 (2.5) | NA | No increase / decrease |
| Gerhards, Mannheim / Germany [44] | Mixed | Mean anti-RBD/S1 (SD), U/ml | Any persistent symptoms | | 22 | 299.3 (548.7) | NA | No increase / decrease |
| Gerhards, Mannheim / Germany [44] | Mixed | Mean anti-RBD/S1 (SD), U/ml | No persistent symptoms | | 25 | 354.6 (408.9) | NA | No increase / decrease |
| Lier, Saxony / Germany [63] | Non-hospitalized | Median (IQR), anti-RBD (S/CO) | Neuropsychiatric phenotype | | 105 | 2,250 (9,897) | NA | Decrease |
| Lier, Saxony / Germany [63] | Non-hospitalized | Median (IQR), anti-RBD (S/CO) | Without phenotype | | 55 | 5,256 (12,518) | NA | Decrease |
| Varnai, Hungary / Pecs [30] | Mixed | Median anti-Spike, IQR, (U/mL) | Severe fatigue | | 57 | 3,723 (10,021) | NA | No increase / decrease |
| Varnai, Hungary / Pecs [30] | Mixed | Median anti-Spike, IQR, (U/mL) | Non-severe fatigue | | 50 | 6,949 (11,070) | NA | No increase / decrease |
| **Serological sampling 9 - < 12 months post COVID-19 (studies = 1; participants = 121)** | | | | | | | | |
| Zhan, Xiangyang / China [56] | Hospitalized | Beta (95% CI), total anti-RBD | Any persistent symptoms vs no persistent symptoms | | 121 | -0.4  (-0.57, -0.13) | Time from discharge, length of stay, age, sex, severe disease, glucocorticoids, interferons | Decrease |
| Zhan, Xiangyang / China [56] | Hospitalized | Beta (95% CI), anti-RBD | Any persistent symptoms vs no persistent symptoms | | 121 | -0.2 (-0.38, -0.05) | Time from discharge, length of stay, age, sex, severe disease, glucocorticoids, interferons | Decrease |
| ^a^Trend as reported by study authors  CSQ - COVID-19 symptoms questionnaire; IQR - interquartile range; RU - relative light units; S/CO - signal to cut-off ratio; NA - Not applicable; NR - Not reported; T1 - Visit 1; T4 - Visit 4; OR - odds ratio; CI - confidence interval | | | | | | | | |

| **Table S6: Neutralizing antibody comparisons among groups with and without persistent symptoms** ≥**12 weeks post COVID-19, stratified by time interval (months) between COVID-19 infection and serological sampling** | | | | | | | | |
| --- | --- | --- | --- | --- | --- | --- | --- | --- |
| **Study cohort** | **LOC** | **Measure / unit** |  | **Comparisons** | | | | **Overall trend^a^** |
|  |  |  | **Group** | | **Participants assessed (N)** | **Results** | **Variables adjusted** |  |
| **Serological sampling < 3 months post COVID-19 (studies = 3; participants = 675)** | | | | | | | | |
| Blomberg, Bergen / Norway [58] | Mixed | Microneutralizing titres RR (95% CI) | Fatigue score  (0-33) | | 293 | 1.1 (1.08, 1.19) | Sex, age, BMI, asthma/COPD, hypertension, chronic heart disease, rheumatic disease, diabetes, severity of acute illness, days in hospital | Increase |
| Blomberg, Bergen / Norway [58] | Mixed | Mean microneutralizing titres (95% CI) | Fatigue | | 108 | 2.2 (2.10, 2.40) | NA | Increase |
| Blomberg, Bergen / Norway [58] | Mixed | Mean microneutralizing titres (95% CI) | No fatigue | | 185 | 1.9 (1.80, 2.00) | NA | Increase |
| Blomberg, Bergen / Norway [58] | Mixed | Microneutralizing titres, RR (95% CI) | Fatigue vs no fatigue | | 293 | 1.8 (1.27, 2.52) | NA | Increase |
| Peluso, San Francisco / California [49] | Mixed | Differences in median neutralizing capacity (infectious dose, 50% [ID50]) at T1 and T4 | Any persistent symptoms vs no persistent symptoms | | 66 | Not specified | NA | No increase / decrease |
| Wynberg, Amsterdam, The Netherlands [55] | Mixed | Neutralizing antibodies, median difference in posterior means (95% credible interval) | Any persistent symptoms vs no persistent symptoms | | 316 | 0.17  (-0.05, 0.40) | NA | No increase / decrease |
| **Serological sampling 3 - < 6 months post COVID-19 (n = 2; participants = 162)** | | | | | | | | |
| Peluso, San Francisco / California [49] | Mixed | Differences in median neutralizing capacity (infectious dose, 50% [ID50]) at T1 and T4 | Any persistent symptoms vs no persistent symptoms | | 66 | Not specified | NA | No increase / decrease |
| Seeßle, Heidelberg / Germany [52] | Mixed | Median relative S1-ACE2 competition efficiency (%): Group differences based on Mann-Whitney U test | Any persistent symptoms vs no persistent symptoms | | 96 | Not specified | NA | No increase / decrease |
| **Serological sampling 6 - < 9 months post COVID-19 (studies = 2; participants = 256)** | | | | | | | | |
| Lier, Saxony / Germany [63] | Non-hospitalized | Median (IQR), (S/CO) Neutralizing antibodies | Neuropsychiatric phenotype | | 105 | 465 (1,398) | NA | Decrease |
| Lier, Saxony / Germany [63] | Non-hospitalized | Median (IQR), (S/CO) Neutralizing antibodies | Without phenotype | | 55 | 746 (1,539) | NA | Decrease |
| Seeßle, Heidelberg / Germany [52] | Mixed | Median relative S1-ACE2 competition efficiency (%): Group differences based on Mann-Whitney U test | Any persistent symptoms vs no persistent symptoms | | 96 | Not specified | NA | No increase / decrease |
| **Serological sampling 9 - 12 months post COVID-19 (studies = 3; participants = 289)** | | | | | | | | |
| Seeßle, Heidelberg / Germany [52] | Mixed | Median relative S1-ACE2 competition efficiency (%): Group differences based on Mann-Whitney U test | Any persistent symptoms vs no persistent symptoms | | 96 | Not specified | NA | No increase / decrease |
| Zhan, Xiangyang / China [56] | Hospitalized | Median 50% pseudovirus neutralization titers (pNT50) | Any persistent symptoms | | 36 | 18.8 | NA | Decrease |
| Zhan, Xiangyang / China [56] | Hospitalized | Median 50% pseudovirus neutralization titers (pNT50) | No persistent symptoms | | 85 | 29.2 | NA | Decrease |
| García-Abellán, Alicante / Spain [28] | Hospitalized | Median (IQR) NeutraLISA,% IH | Any persistent symptoms | | 14 | 27.3 (59.0) | NA | No increase / decrease |
| García-Abellán, Alicante / Spain [28] | Hospitalized | Median (IQR) NeutraLISA,% IH | No persistent symptoms | | 58 | 69.7 (44.0) | NA | No increase / decrease |
| García-Abellán, Alicante / Spain [28] | Hospitalized | NeutraLISA positive,  N (%) | Any persistent symptoms | | 14 | 6.0 (42.9) | NA | Decrease |
| García-Abellán, Alicante / Spain [28] | Hospitalized | NeutraLISA positive,  N (%) | No persistent symptoms | | 58 | 46.0 (79.3) | NA | Decrease |
| García-Abellán, Alicante / Spain [28] | Hospitalized | aHR (95% CI) NeutraLISA positive, AU/Ml | Any persistent symptoms vs no persistent symptoms | | 72 | 0.98  (0.97, 0.99) | Sex and ICU stay | Decrease |
| ^a^Trend as reported by study authors  CSQ - COVID-19 symptoms questionnaire; IQR - interquartile range; RU - relative light units; S/CO - signal to cut-off ratio; NA - Not applicable; NR - Not reported; T1 - Visit 1; T4 - Visit 4; OR - odds ratio; CI - confidence interval; IH – inhibition; aHR – adjusted hazard ratio; RR - risk ratio | | | | | | | | |

**Search strategy - OVID Medline and Embase databases searched October 22, 2022**

**OVID Medline**

1	(longcovid* or long covid* or longcoronavirus* or longcorona* virus* or long coronavirus* or long corona* virus* or longcorono* virus* or long coronovirus* or long corono* virus* or longcoronavirinae* or longcorona* virinae* or long coronavirinae* or long corona* virinae* or longCov or long Cov or longsars* or long sars* or "long severe acute respiratory syndrome*" or longncov* or long ncov* or longhcov* or long hcov*).ti,ab,kf.	1759

2	((long or longterm or long term or longhaul or long haul or post acute or postacute or after acute or sequela* or protracted or post-infect* or post-viral or post-discharg* or non-recover* or nonrecover* or PCC) adj7 (COVID* or coronavirus* or corona virus* or 2019-nCoV or 19nCoV or 2019nCoV or nCoV or n-CoV or SARS-CoV-2 or SARS-CoV2 or SARSCoV-2 or SARSCoV2 or 2019-novel CoV or Sars-coronavirus2 or novel CoV)).ti,ab,kf.	7622

3	((chronic or persist* or linger* or continuing or continual) adj3 (COVID* or coronavirus* or corona virus* or 2019-nCoV or 19nCoV or 2019nCoV or nCoV or n-CoV or SARS-CoV-2 or SARS-CoV2 or SARSCoV-2 or SARSCoV2 or 2019-novel CoV or Sars-coronavirus2 or novel CoV)).ti,ab,kf.	2005

4	(post covid* or postcovid* or post corona* or postcorona* or ((post* or survivor*) adj3 (COVID* or coronavirus* or corona virus* or 2019-nCoV or 19nCoV or 2019nCoV or nCoV or n-CoV or SARS-CoV-2 or SARS-CoV2 or SARSCoV-2 or SARSCoV2 or 2019-novel CoV or Sars-coronavirus2 or novel CoV))).ti,ab,kf.	8624

5	1 or 2 or 3 or 4	15750

6	limit 5 to yr="2020 -Current"	14884

7	Epidemiologic Methods/ or exp Epidemiologic Studies/ or Observational Studies as Topic/ or Clinical Studies as Topic/ or single-case studies as topic/ or (Observational Study or Validation Studies or Clinical Study).pt. or cohort*.ti,ab,kf. or (prospective adj7 (study or studies or design or analysis or analyses)).ti,ab,kf. or ((follow up or followup) adj7 (study or studies or design or analysis or analyses)).ti,ab,kf. or ((longitudinal or longterm or (long adj term)) adj7 (study or studies or design or analysis or analyses or data)).ti,ab,kf. or (retrospective adj7 (study or studies or design or analysis or analyses or data or review)).ti,ab,kf. or ((case adj control) or (case adj comparison) or (case adj controlled)).ti,ab,kf. or (case-referent adj3 (study or studies or design or analysis or analyses)).ti,ab,kf. or (population adj3 (study or studies or analysis or analyses)).ti,ab,kf. or (descriptive adj3 (study or studies or design or analysis or analyses)).ti,ab,kf. or ((multidimensional or (multi adj dimensional)) adj3 (study or studies or design or analysis or analyses)).ti,ab,kf. or (cross adj sectional adj7 (study or studies or design or research or analysis or analyses or survey or findings)).ti,ab,kf. or ((natural adj experiment) or (natural adj experiments)).ti,ab,kf. or (quasi adj (experiment or experiments or experimental)).ti,ab,kf. or ((non experiment or nonexperiment or non experimental or nonexperimental) adj3 (study or studies or design or analysis or analyses)).ti,ab,kf. or (prevalence adj3 (study or studies or analysis or analyses)).ti,ab,kf. or case series.ti,ab,kf.	4083612

8	6 and 7	4760

9	editorial/ or letter/ or case reports/ or clinical conference/ or exp Computer Simulation/ or Molecular Dynamics Simulation/ or case study.ti,ab,kf.	4221680

10	8 not 9	4469

**Embase**

1	(longcovid* or long covid* or longcoronavirus* or longcorona* virus* or long coronavirus* or long corona* virus* or longcorono* virus* or long coronovirus* or long corono* virus* or longcoronavirinae* or longcorona* virinae* or long coronavirinae* or long corona* virinae* or longCov or long Cov or longsars* or long sars* or "long severe acute respiratory syndrome*" or longncov* or long ncov* or longhcov* or long hcov*).ti,ab,kf.	2310

2	((long or longterm or long term or longhaul or long haul or post acute or postacute or after acute or sequela* or protracted or post-infect* or post-viral or post-discharg* or non-recover* or nonrecover* or PCC) adj7 (COVID* or coronavirus* or corona virus* or 2019-nCoV or 19nCoV or 2019nCoV or nCoV or n-CoV or SARS-CoV-2 or SARS-CoV2 or SARSCoV-2 or SARSCoV2 or 2019-novel CoV or Sars-coronavirus2 or novel CoV)).ti,ab,kf.	9948

3	((chronic or persist* or linger* or continuing or continual) adj3 (COVID* or coronavirus* or corona virus* or 2019-nCoV or 19nCoV or 2019nCoV or nCoV or n-CoV or SARS-CoV-2 or SARS-CoV2 or SARSCoV-2 or SARSCoV2 or 2019-novel CoV or Sars-coronavirus2 or novel CoV)).ti,ab,kf.	2651

4	(post covid* or postcovid* or post corona* or postcorona* or ((post* or survivor*) adj3 (COVID* or coronavirus* or corona virus* or 2019-nCoV or 19nCoV or 2019nCoV or nCoV or n-CoV or SARS-CoV-2 or SARS-CoV2 or SARSCoV-2 or SARSCoV2 or 2019-novel CoV or Sars-coronavirus2 or novel CoV))).ti,ab,kf.	12424

5	long COVID/	2436

6	1 or 2 or 3 or 4 or 5	21607

7	observational study/	292348

8	cohort analysis/	909623

9	longitudinal study/	180803

10	follow up/	1952398

11	retrospective study/	1330625

12	exp case control study/	212493

13	exp case control study/	212493

14	quasi experimental study/	10089

15	prospective study/	805754

16	(observational adj3 (study or studies or design or analysis or analyses)).ti,ab,kf.	314519

17	cohort*.ti,ab,kf.	1340888

18	(prospective adj7 (study or studies or design or analysis or analyses)).ti,ab,kf.	765315

19	((follow up or followup) adj7 (study or studies or design or analysis or analyses)).ti,ab,kf.	261348

20	((longitudinal or longterm or (long adj term)) adj7 (study or studies or design or analysis or analyses or data)).ti,ab,kf.	466096

21	(retrospective adj7 (study or studies or design or analysis or analyses or data or review)).ti,ab,kf.	1056842

22	((case adj control) or (case adj comparison) or (case adj controlled)).ti,ab,kf.	201859

23	(case-referent adj3 (study or studies or design or analysis or analyses)).ti,ab,kf.	694

24	(population adj3 (study or studies or analysis or analyses)).ti,ab,kf.	331742

25	(descriptive adj3 (study or studies or design or analysis or analyses)).ti,ab,kf.	153133

26	((multidimensional or (multi adj dimensional)) adj3 (study or studies or design or analysis or analyses)).ti,ab,kf.	5462

27	(cross adj sectional adj7 (study or studies or design or research or analysis or analyses or survey or findings)).ti,ab,kf.	523781

28	((natural adj experiment) or (natural adj experiments)).ti,ab,kf.	3246

29	(quasi adj (experiment or experiments or experimental)).ti,ab,kf.	23118

30	((non experiment or nonexperiment or non experimental or nonexperimental) adj3 (study or studies or design or analysis or analyses)).ti,ab,kf.	2200

31	(prevalence adj3 (study or studies or analysis or analyses)).ti,ab,kf.	67711

32	case series.ti,ab,kf.	136927

33	7 or 8 or 9 or 10 or 11 or 12 or 13 or 14 or 15 or 16 or 17 or 18 or 19 or 20 or 21 or 22 or 23 or 24 or 25 or 26 or 27 or 28 or 29 or 30 or 31 or 32	5982094

34	6 and 33	9531

35	limit 34 to yr="2020 -Current"	9078

36	editorial/ or letter/ or case reports/ or clinical conference/ or exp Computer Simulation/ or Molecular Dynamics Simulation/ or case study.ti,ab,kf.	2278867

37	35 not 36	8757

38	limit 37 to "remove medline records"	4471

| 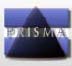  **PRISMA 2020 Checklist** | | | |
| --- | --- | --- | --- |
| **Section and Topic** | **Item #** | **Checklist item** | **Location where item is reported** |
| **TITLE** | | |  |
| Title | 1 | Identify the report as a systematic review. | 1 |
| **ABSTRACT** | | |  |
| Abstract | 2 | See the PRISMA 2020 for Abstracts checklist. | 2 |
| **INTRODUCTION** | | |  |
| Rationale | 3 | Describe the rationale for the review in the context of existing knowledge. | 3 - 4 |
| Objectives | 4 | Provide an explicit statement of the objective(s) or question(s) the review addresses. | 4 |
| **METHODS** | | |  |
| Eligibility criteria | 5 | Specify the inclusion and exclusion criteria for the review and how studies were grouped for the syntheses. | 4,6 |
| Information sources | 6 | Specify all databases, registers, websites, organisations, reference lists and other sources searched or consulted to identify studies. Specify the date when each source was last searched or consulted. | 4 |
| Search strategy | 7 | Present the full search strategies for all databases, registers and websites, including any filters and limits used. | 4, Supplement |
| Selection process | 8 | Specify the methods used to decide whether a study met the inclusion criteria of the review, including how many reviewers screened each record and each report retrieved, whether they worked independently, and if applicable, details of automation tools used in the process. | 5 |
| Data collection process | 9 | Specify the methods used to collect data from reports, including how many reviewers collected data from each report, whether they worked independently, any processes for obtaining or confirming data from study investigators, and if applicable, details of automation tools used in the process. | 5 |
| Data items | 10a | List and define all outcomes for which data were sought. Specify whether all results that were compatible with each outcome domain in each study were sought (e.g. for all measures, time points, analyses), and if not, the methods used to decide which results to collect. | 5,6 |
|  | 10b | List and define all other variables for which data were sought (e.g. participant and intervention characteristics, funding sources). Describe any assumptions made about any missing or unclear information. | 5,6 |
| Study risk of bias assessment | 11 | Specify the methods used to assess risk of bias in the included studies, including details of the tool(s) used, how many reviewers assessed each study and whether they worked independently, and if applicable, details of automation tools used in the process. | 5,6 |
| Effect measures | 12 | Specify for each outcome the effect measure(s) (e.g. risk ratio, mean difference) used in the synthesis or presentation of results. | 6 |
| Synthesis methods | 13a | Describe the processes used to decide which studies were eligible for each synthesis (e.g. tabulating the study intervention characteristics and comparing against the planned groups for each synthesis (item #5)). | 5,6 |
|  | 13b | Describe any methods required to prepare the data for presentation or synthesis, such as handling of missing summary statistics, or data conversions. | 6 |
|  | 13c | Describe any methods used to tabulate or visually display results of individual studies and syntheses. | 6 |
|  | 13d | Describe any methods used to synthesize results and provide a rationale for the choice(s). If meta-analysis was performed, describe the model(s), method(s) to identify the presence and extent of statistical heterogeneity, and software package(s) used. | 6 |
|  | 13e | Describe any methods used to explore possible causes of heterogeneity among study results (e.g., subgroup analysis, meta-regression). | 5,6 |
|  | 13f | Describe any sensitivity analyses conducted to assess robustness of the synthesized results. | NA |
| Reporting bias assessment | 14 | Describe any methods used to assess risk of bias due to missing results in a synthesis (arising from reporting biases). | 5,6 |
| Certainty assessment | 15 | Describe any methods used to assess certainty (or confidence) in the body of evidence for an outcome. | 5,6 |
| **RESULTS** | | |  |
| Study selection | 16a | Describe the results of the search and selection process, from the number of records identified in the search to the number of studies included in the review, ideally using a flow diagram. | 6, Fig 1 |
|  | 16b | Cite studies that might appear to meet the inclusion criteria, but which were excluded, and explain why they were excluded. | 6 |
| Study characteristics | 17 | Cite each included study and present its characteristics. | Table 1 |
| Risk of bias in studies | 18 | Present assessments of risk of bias for each included study. | 7, Tables 2 - 4 |
| Results of individual studies | 19 | For all outcomes, present, for each study: (a) summary statistics for each group (where appropriate) and (b) an effect estimate and its precision (e.g. confidence/credible interval), ideally using structured tables or plots. | 8,9, Tables S1 - S6 |
| Results of syntheses | 20a | For each synthesis, briefly summarise the characteristics and risk of bias among contributing studies. | Tables 1 - 4 |
|  | 20b | Present results of all statistical syntheses conducted. If meta-analysis was done, present for each the summary estimate and its precision (e.g. confidence/credible interval) and measures of statistical heterogeneity. If comparing groups, describe the direction of the effect. | 8,9, Tables S1 - S6 |
|  | 20c | Present results of all investigations of possible causes of heterogeneity among study results. | 9, 11 - 13 |
|  | 20d | Present results of all sensitivity analyses conducted to assess the robustness of the synthesized results. | NA |
| Reporting biases | 21 | Present assessments of risk of bias due to missing results (arising from reporting biases) for each synthesis assessed. | 6, Tables 2 - 4 |
| Certainty of evidence | 22 | Present assessments of certainty (or confidence) in the body of evidence for each outcome assessed. | 10,11 |
| **DISCUSSION** | | |  |
| Discussion | 23a | Provide a general interpretation of the results in the context of other evidence. | 10,11 |
|  | 23b | Discuss any limitations of the evidence included in the review. | 10 - 13 |
|  | 23c | Discuss any limitations of the review processes used. | 13 |
|  | 23d | Discuss implications of the results for practice, policy, and future research. | 10,14 |
| **OTHER INFORMATION** | | |  |
| Registration and protocol | 24a | Provide registration information for the review, including register name and registration number, or state that the review was not registered. | 4 |
|  | 24b | Indicate where the review protocol can be accessed, or state that a protocol was not prepared. | 4 |
|  | 24c | Describe and explain any amendments to information provided at registration or in the protocol. | NA |
| Support | 25 | Describe sources of financial or non-financial support for the review, and the role of the funders or sponsors in the review. | 14 |
| Competing interests | 26 | Declare any competing interests of review authors. | 14 |
| Availability of data, code and other materials | 27 | Report which of the following are publicly available and where they can be found: template data collection forms; data extracted from included studies; data used for all analyses; analytic code; any other materials used in the review. | 14 |
| *From:* Page MJ, McKenzie JE, Bossuyt PM, Boutron I, Hoffmann TC, Mulrow CD, et al. The PRISMA 2020 statement: an updated guideline for reporting systematic reviews. BMJ 2021;372:n71. doi: 10.1136/bmj.n71 | | | |
